# Respiratory and Gut Microbiota Correlate with Lung Function Recovery after Severe COVID-19

**DOI:** 10.64898/2026.02.09.26345630

**Authors:** Sabine V Stadler, Luca C Stickley, Eric Bernasconi, Seher Guney, Aurélien Trompette, Lise Piquilloud, Manuela Funke-Chambour, Christophe von Garnier, Niki D Ubags

## Abstract

**Rationale:** Severe SARS-CoV-2 infection induces disrupted oropharyngeal and gut microbiota during acute disease which may persist and contribute to the development of post-acute pulmonary sequelae. To date, it is unclear whether dysbiosis following severe disease is linked to long-term pulmonary function impairment.

**Objectives:** To determine associations between oropharyngeal and gut microbiota composition with lung function after severe COVID-19.

**Methods:** 16S and internal transcribed spacer (ITS) rRNA amplicon sequencing were performed on oropharyngeal (16S and ITS) and rectal (16S) swabs at 3-, 6- and 12-months post-hospitalisation from 83 subjects previously admitted to the ICU with severe COVID-19 (Swiss COVIDlung study, NCT04581135). Subjects underwent 1-3 follow-up visits during which lung function testing was performed to investigate associations with microbiota composition.

**Measurements and Main Results:** The oropharyngeal microbiota of subjects having suffered from COVID-19-related-severe acute non-cardiogenic hypoxemic respiratory failure with bilateral lung infiltrates (AHRF-BLI) was characterized by decreased α-diversity and the presence of differentially abundant taxa. Subjects who recovered in lung function (TLC, FVC, FEV_1_ and DLCO >Lower Limit of Normal) had a distinct oropharyngeal and gut microbiota composition compared to those whose lung function never recovered. Fungal analysis of oropharyngeal samples revealed the presence of three distinct clusters which were characterized by distinct lung-function associated bacterial-fungal co-occurrence.

**Conclusions:** This study provide first insights into the role of the airway and gut microbiota in the development of long-term pulmonary sequelae after severe SARS-CoV-2 infection, shedding the light on the potential of the microbiota for preventive and therapeutic strategies in severe COVID-19.

## Introduction

The Coronavirus disease 2019 (COVID-19) pandemic, caused by the novel coronavirus SARS-CoV-2, has impacted over 770 million people globally(1). SARS-CoV-2 primarily targets the respiratory system, with clinical presentations ranging from asymptomatic to severe such as acute respiratory distress syndrome (ARDS)(2,3). Currently, enduring health consequences among COVID-19 survivors are increasingly recognized, raising international concern regarding the disease burden. COVID-19-induced impairments in diffusion capacity for carbon monoxide (DLCO), total lung capacity (TLC), and forced vital capacity (FVC) often persist for 12 months in critically ill patients(4–6), strongly motivating further investigation into how severe SARS-CoV-2 infection leads to long-term respiratory impairment.

Viral lung infections can disrupt the respiratory tract microbiota(7–10), and such dysbiosis can induce inflammation, tissue destruction and remodelling(11–13), thereby affecting lung function(14). Although earlier studies have shown disturbed oropharyngeal microbiota in acute COVID-19(15–17), associations with long-term pulmonary sequelae remain unclear. Given the risk for fungal pulmonary infections in severe COVID-19, respiratory tract mycobiota is an important factor to consider (18). Additionally, the gut microbiota modifies lung immunity and respiratory disease development via the gut-lung axis(19,20). The close relationship between these compartments is reinforced by findings of gut-associated bacteria enrichment in the lung microbiota of patients with acute ARDS(21). Although acute COVID-19 can alter the gut microbiota (22,23), gut-lung crosstalk and correlations with lung function in this context remain unexplored.

This study provides evidence of associations between lung function and alterations in oropharyngeal and gut microbiomes up to 12 months post severe COVID-19. Furthermore, bacteria found in both compartments negatively correlate with lung function. Finally, we describe novel insights into three distinct mycobiota profiles and their inter-kingdom interactions related to post-COVID-19 lung function recovery.

## Materials and Methods

### Study design and data collection

Subjects were prospectively recruited from May 1, 2020 to December 31, 2021 as part of the Swiss COVID-19 lung study, a multicentre prospective observational cohort investigating long-term pulmonary sequelae of SARS-CoV-2 infection. The study was approved by the Swiss Ethics Committees on research involving humans (KEK 2020-00799, Identifier NCT04581135)(24). Inclusion and exclusion criteria of the national study have previously been reported(24), and all patients signed written informed consent.

Patients included in the current analyses (n=83) suffered from severe COVID-19, as defined by ICU admission (Lausanne University Hospital, CHUV), with or without severe acute non-cardiogenic hypoxemic respiratory failure with bilateral lung infiltrates (AHFR-BLI). The latter was defined as acute hypoxemia with PaO₂/FiO₂ ≤ 200 mmHg requiring invasive or non-invasive ventilatory support with oxygen supplementation, associated with bilateral pulmonary infiltrates on chest X-ray or CT scan without evidence of cardiogenic pulmonary edema(3).

Data and specimens were collected during follow-up study visits at 3-, 6- and 12-months after hospitalisation, including pulmonary function tests (TLC, FVC, Forced Expiratory Volume in 1s (FEV_1_), and DLCO), as well as oropharyngeal and stool swabs for microbial DNA isolation (**Figure 1A**). Lung function normality thresholds were determined using the latest ERS/ATS standards (25). For each time-point, a z-score value < 1.645 (lower limit) was considered abnormal.

**Figure 1.**
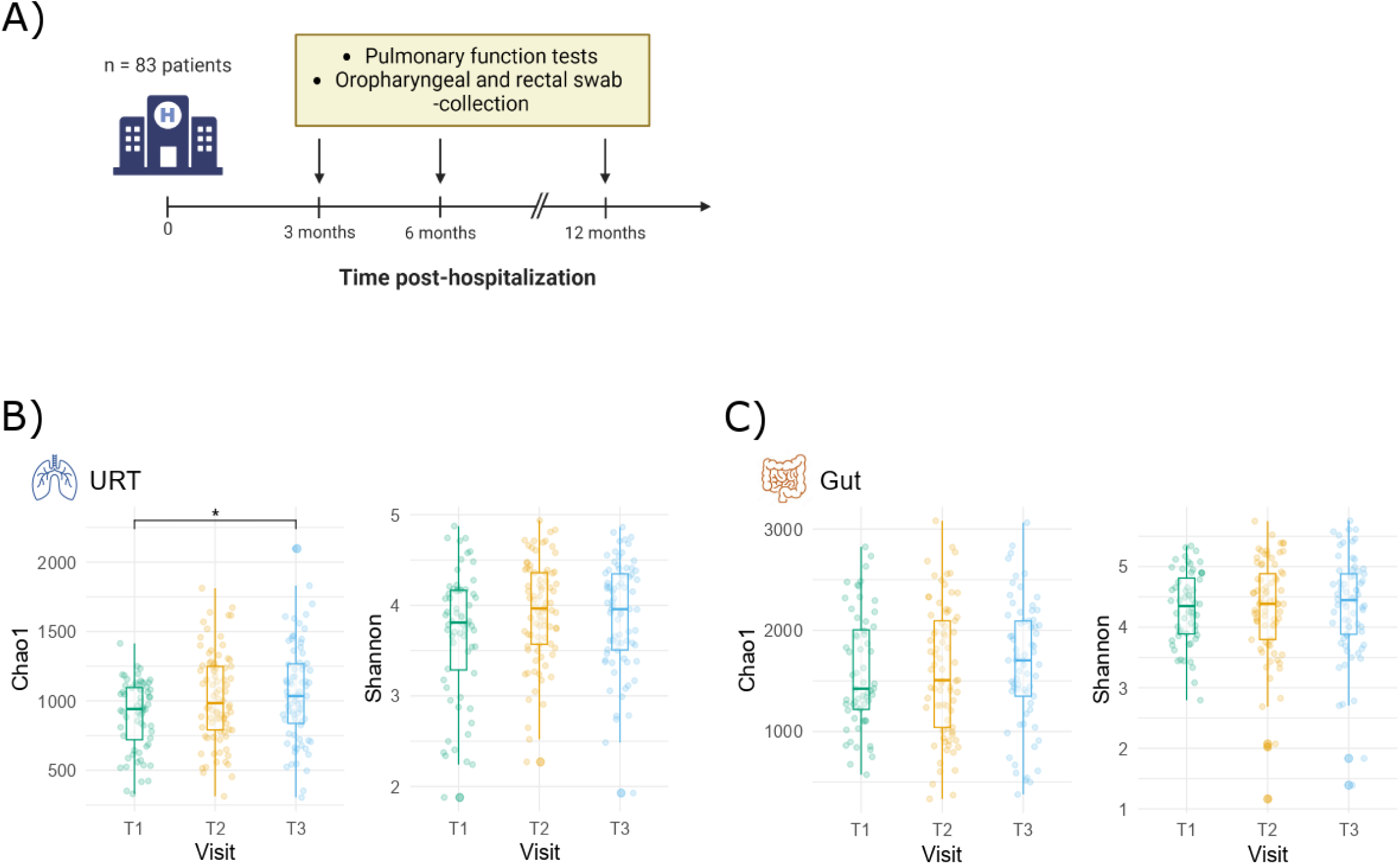
– Study design and URT and gut bacterial alpha-diversity. (**A**) Study design. Patients (n = 83) were followed-up at 3- (T1), 6- (T2) and 12- (T3) months after hospitalization for severe COVID-19. Collected data and specimens included pulmonary function tests (TLC, DLCO, FVC and FEV_1_), as well as oropharyngeal and rectal swabs. (**B**) Alpha diversity dynamics (Chao1 and Shannon) in the URT over the study period. Each dot represents one sample. T1 = 3-months, T2 = 6-months, T3 = 12-months post-hospitalization. Comparisons were made using Kruskal-Wallis with Dunn’s post-hoc test and Bonferroni adjustment. *P-value significance: *. < 0.05, **. ≤ 0.01, ***. ≤ 0.001.* (**C**) Alpha diversity dynamics (Chao1 and Shannon) in the gut over the study period. Each dot represents one sample. T1 = 3-months, T2 = 6-months, T3 = 12-months post-hospitalization. Comparisons were made using Kruskal-Wallis with Dunn’s post-hoc test and Bonferroni adjustment. *P-value significance: *. < 0.05, **. ≤ 0.01, ***. ≤ 0.001*.

### Bacterial 16S rRNA gene and fungal ITS amplicon sequencing

The 16S rRNA gene was amplified by a 1-step PCR targeting the V1-V2 hypervariable region using universal primers F-27 and R-338. The ITS region was amplified by a 2-step PCR using 8 forward and 7 reverse primers(26,27) (**Supplementary Table E1**). Sequencing was performed using the Illumina MiSeq v2 platform (2 x 250bp) (Lausanne Genomic Technologies Facility, University of Lausanne, Switzerland). Additional details in online supplements.

### Bioinformatics analysis

Demultiplexed sequences were filtered, trimmed, denoised, merged, chimeras removed, and assigned taxonomies using the Divisive Amplicon Denoising Algorithm 2 (DADA2) package (version 1.32.0), in R (v4.4.0) for Linux(28). Filtering parameters were determined using FIGARO(29). Cutadapt primer removal was used on ITS reads(30). The prevalence method of the decontam package (v1.24.0)(31) identified and removed 81, 55, and 1 contaminant taxa from the oropharyngeal 16S, gut 16S, and oropharyngeal ITS datasets, respectively.

### Graphical Analysis

Graphs were produced using SpiecEasi(32,33) and further analysed with iGraph(34). SPIEC-EASI was run with the meinshausen-buhlmann method, nlambda of 50, lambda.min.ratio of 0.01, and 99 repetitions. Amplicon Sequence Variants (ASVs) with at least one read in 50% of samples were used as input.

### Statistical analysis

Wilcoxon test was used for single group comparisons. Multiple group comparisons were performed using the Kruskal-Wallis test with Dunn’s post-hoc test and Bonferroni’s adjustment. Differences between bacterial communities were tested using permutational multivariate analysis of variance using distance matrices (PERMANOVA). Differences were considered significant at *p* < 0.05. DESeq2 was used to determine differentially abundant taxa between two groups. Unsupervised clustering was performed using the Partitioning Around Medoids K-means (pam) from cluster (version 2.1.6)(35). Mixed effect models were performed using the lme4 package n patients with at least two timepoints(36). R scripts are available at https://github.com/CHUVpulmonology/COVIDlung_microbiota.git.

## Results

### Study population

The study included 83 subjects who suffered from severe COVID-19 and were hospitalised at the Intensive Care Unit (ICU) of the Lausanne University Hospital (CHUV) between January and December 2020. Subjects were followed-up at 3- (T1), 6- (T2) and 12- (T3) months post-hospitalisation (**Figure 1A**). In total, subjects were predominantly male (73%) with a high BMI (average 29 kg/m^2^). Forty-nine subjects (60%) required intubation, and 61 (73%) received antibiotic treatment. Sixty-one subjects (73%) suffered from COVID-19-related AHFR-BLI during their hospitalisation (**Table 1**). Overall, patient characteristics remained homogeneous between time-points (**Table 1**). Sample availability per subject is presented in **Figure E1**.

**Table 1.**
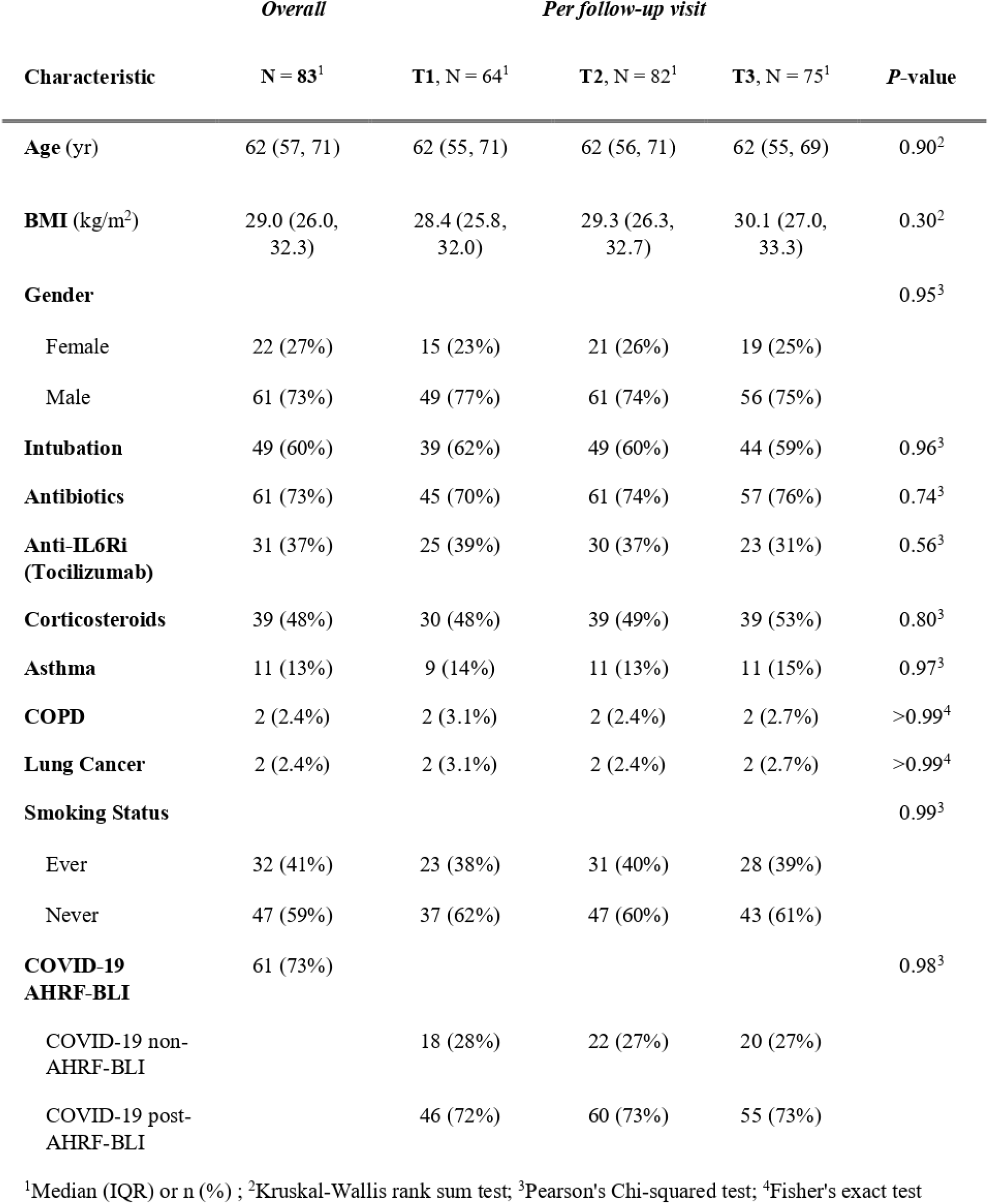
Demographics table of the overall patient cohort (n = number of subjects) and per follow-up visit (n = number of samples). T1 = 3-months, T2 = 6 -months, T3 = 12 -months post-hospitalization.

### Respiratory function and microbiota α-diversity in the URT and gut over time

To characterize long-term changes in microbiota composition and lung function following severe COVID-19, we examined temporal patterns of microbiota diversity and pulmonary function parameters (TLC, FVC, FEV_1_ and DLCO) in all study subjects (n=83). Long-term respiratory recovery was assessed using a longitudinal mixed-effects linear model with a per patient random intercept. For all measured lung function metrics, we found T2 and T3 follow-up visits to be statistically significant as compared to T1 (**Table 2**). In all cases, overall lung function metrics increased at T2 and T3. Recovery was independent of corticosteroid and antibiotic administration during hospitalisation (**Table 2**).

**Table 2.** Fixed effects estimates from a mixed-effects model of Forced Vital Capacity (FVC), Forced Expiratory Volume in 1 second (FEV_1_), Total Lung Capacity (TLC), and Diffusing capacity of the Lungs for Carbon monoxide (DLCO) z-score by timepoint and medication.

| Independent Variable | Coefficient (95% CI) | Std. Error | Statistic | df | P-value |
| --- | --- | --- | --- | --- | --- |
| **FVC** |  |  |  |  |  |
| (Intercept) | -1.162 (-1.967 to -0.357) | 0.399 | -2.912 | 43.051 | 0.006 |
| T2 | 0.44 (0.191 to 0.689) | 0.125 | 3.528 | 62.168 | 0.001 |
| T3 | 0.587 (0.324 to 0.85) | 0.132 | 4.464 | 62.693 | <0.001 |
| Antibiotics (Yes) | 0.062 (-0.749 to 0.874) | 0.401 | 0.156 | 39.469 | 0.877 |
| Corticosteroids (Yes) | -0.357 (-1.045 to 0.331) | 0.340 | -1.049 | 39.680 | 0.300 |
| Inhaled Corticosteroids (Yes) | 0.077 (-2.241 to 2.395) | 1.148 | 0.067 | 40.824 | 0.947 |
| **FEV<sub>1</sub>** |  |  |  |  |  |
| (Intercept) | -1.004 (-1.794 to -0.215) | 0.392 | -2.565 | 42.836 | 0.014 |
| T2 | 0.392 (0.166 to 0.619) | 0.113 | 3.462 | 62.216 | 0.001 |
| T3 | 0.564 (0.325 to 0.803) | 0.120 | 4.719 | 62.663 | <0.001 |
| Antibiotics (Yes) | 0.209 (-0.59 to 1.007) | 0.395 | 0.528 | 39.769 | 0.600 |
| Corticosteroids (Yes) | -0.231 (-0.908 to 0.446) | 0.335 | -0.689 | 39.950 | 0.495 |
| Inhaled Corticosteroids (Yes) | -0.55 (-2.829 to 1.729) | 1.128 | -0.487 | 40.920 | 0.629 |
| **TLC** |  |  |  |  |  |
| (Intercept) | -2.182 (-2.961 to -1.404) | 0.384 | -5.676 | 37.909 | 0.000 |
| T2 | 0.581 (0.388 to 0.775) | 0.097 | 6.017 | 54.717 | 0.000 |
| T3 | 0.822 (0.608 to 1.036) | 0.107 | 7.706 | 55.297 | 0.000 |
| Antibiotics (Yes) | 0.005 (-0.791 to 0.8) | 0.392 | 0.012 | 35.864 | 0.990 |
| Corticosteroids (Yes) | -0.189 (-0.885 to 0.506) | 0.343 | -0.552 | 36.011 | 0.584 |
| Inhaled Corticosteroids (Yes) | 0.336 (-1.905 to 2.577) | 1.106 | 0.304 | 36.575 | 0.763 |
| **DLCO** |  |  |  |  |  |
| (Intercept) | -2.477 (-2.974 to -1.981) | 0.247 | -10.035 | 48.122 | 0.000 |
| T2 | 0.224 (0.002 to 0.446) | 0.111 | 2.011 | 64.268 | 0.049 |
| T3 | 0.691 (0.453 to 0.929) | 0.119 | 5.798 | 66.368 | 0.000 |
| Antibiotics (Yes) | 0.113 (-0.366 to 0.592) | 0.237 | 0.476 | 39.239 | 0.636 |
| Corticosteroids (Yes) | 0.206 (-0.195 to 0.607) | 0.198 | 1.041 | 40.038 | 0.304 |
| Inhaled Corticosteroids (Yes) | 0.696 (-0.284 to 1.676) | 0.486 | 1.432 | 42.652 | 0.160 |

To assess microbiota dynamics, we measured α-diversity using Chao1 and Shannon indexes. Bacterial richness in the upper respiratory tract (URT) increased between T1 and T3 (Chao1, *P* = 0.027), whereas it remained stable in the gut (**Figure 1B and C**). Shannon diversity and bacterial load in both the URT and gut remained stable (**Figure 1B and C, Figure E2A and B**). Overall, these findings suggest distinct microbiota dynamics in the respiratory tract and gut during recovery following severe COVID-19.

### COVID-19-related AHFR-BLI induces long-term URT and gut microbiota alterations

Individuals with AHFR-BLI (post-AHFR-BLI) were characterised by a higher BMI, an increased incidence of intubation, and they received more antibiotics and corticosteroids during hospitalization compared to non-AHFR-BLI subjects (**Table E2**). Temporal lung function metrics did not differ between non-AHFR-BLI and post-AHFR-BLI subjects (**Figure E3**). URT α-diversity (Chao1) was significantly reduced in post-AHFR-BLI subjects at all three time-points (T1 (*P* = 0.0072), T2 (*P* = 0.0034) and T3 (*P* = 0.0004)) compared to non-AHFR-BLI subjects (**Figure 2A**). In addition, URT bacterial diversity progressively increased over time in non-AHFR-BLI subjects (T1 to T3 Chao1, *P* = 0.021), whereas post-AHFR-BLI subjects remained at a constant lower diversity level. Interestingly, AHFR-BLI status did not alter gut bacterial diversity (**Figure 2B**) or bacterial load in the URT and gut (**Figure E4A**).

**Figure 2.**
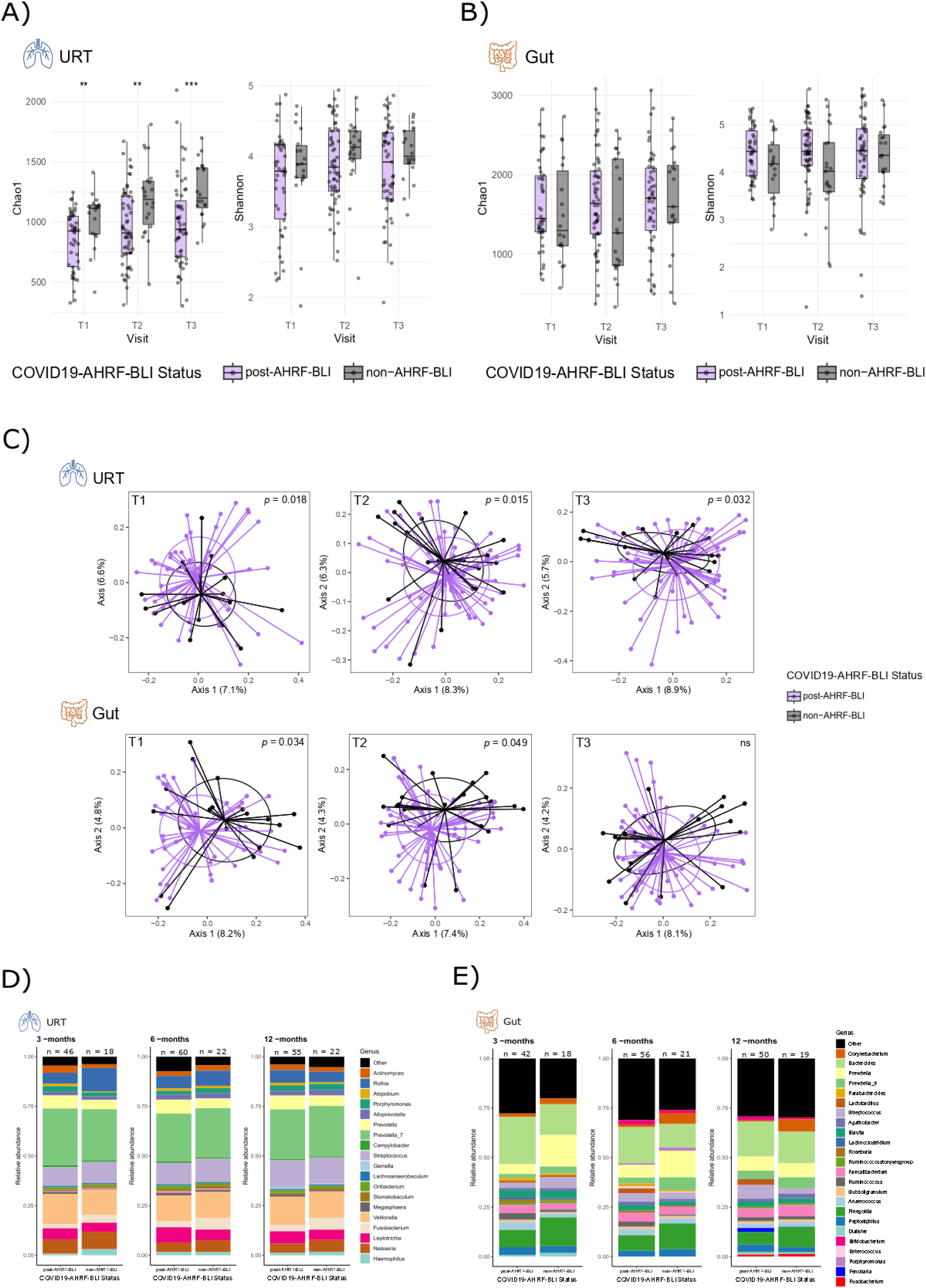
– URT and gut bacterial composition relative to COVID-19-related-AHRF-BLI-status. (**A**) Alpha diversity dynamics (Chao1 and Shannon) in the URT over the study period, relative to COVID-19-related-AHRF-BLIstatus. Each dot represents one sample. T1 = 3-months, T2 = 6-months, T3 = 12-months post-hospitalization. Purple = post-AHRF-BLI, black = non-AHRF-BLI. Comparisons were made using Wilcoxon-test*. P-value significance: *. < 0.05, **. ≤ 0.01, ***. ≤ 0.001.* (**B**) Alpha diversity dynamics (Chao1 and Shannon) in the gut over the study period, relative to COVID-19-related-AHRF-BLI status. Each dot represents one sample. T1 = 3-months, T2 = 6-months, T3 = 12 -months post-hospitalization. Purple = post-AHRF-BLI, black = non-AHRF-BLI. Comparisons were made using Wilcoxon-test. *P-value significance: *. < 0.05, **. ≤ 0.01, ***. ≤ 0.001.* (**C**) Results of the principal coordinate analysis (PCoA) representing differences in beta-diversity (Bray-Curtis) between post-COVID-19 – related-AHF-BLI and non-AHRF-BLI subjects. Each dot represents one sample and each quadrant represents one follow-up visit (T1 = 3-months, T2 = 6-months, T3 = 12-months post-hospitalization). Purple = post-AHRF-BLI, black = non-AHRF-BLI. Axis 1 and 2 indicate the proportion of variance. URT (top) and gut (bottom). Comparisons were made using adonis. (**D**) Relative abundance of the top 20 most abundant genera in the URT at each follow-up visit, per AHRF-BLI-status. The “other” category summarizes all remaining genera. N = number of samples per group. (**E**) Relative abundance of the top 20 most abundant genera in the gut at each follow-up visit, per AHRF-BLI-status. The “other” category summarizes all remaining genera. N = number of samples per group.

Although antibiotic therapy was associated with a decreased URT bacterial diversity at 3-months post-hospitalisation (Chao1, *P* = 0.00007 and Shannon, *P* = 0.013) (**Figure E4B**), this difference was no longer present at later timepoints. In the gut, we observed an overall increase in alpha diversity at T2 in subjects having received any antibiotics during hospitalisation (**Figure E4B**). Moreover, although subjects intubated during hospitalisation had a decreased URT alpha-diversity at 3- and 6- months post- hospitalisation (**Figure E4C**), this finding was independent of AHFR-BLI status (intubated AHFR-BLI subjects (n=48) vs. non-intubated AHFR-BLI subjects (n=13)) (**Figure E4D**). Finally, history of invasive ventilation did not affect gut bacterial diversity (**Figure E4E**).

Assessment of β-diversity (Bray-Curtis distance visualized by PCoA) revealed that URT bacterial composition was significantly different between post-AHFR-BLI and non-AHFR-BLI subjects at all three follow-up visits (**Figure 2C**, (T1) *P* = 0.018, (T2) *P* = 0.015, (T3) *P* = 0.032). In the gut, we only observed a significant difference at T1 and T2 (**Figure 2C**, (T1) *P* = 0.034, (T2) *P* = 0.049), suggesting that AHFR-BLI-induced alterations in gut bacterial community composition recover over time. Although intubation and antibiotics have been described to alter lung microbiota(37), we observed only short-term changes in URT and gut microbiota composition (**Figure E4** and **Table E3**, underscoring the effect of AHFR-BLI on community composition.

Relative abundance analyses demonstrated that *Leptotrichia* was transiently elevated in the URT in post-AHFR-BLI subjects, with consistently higher abundance of *Prevotella* throughout the study period (**Figure 2D**), whereas *Fusobacterium* and *Neisseria* dominated at all follow-up visits in the non- AHFR-BLI group. In the gut, *Bacteroides*, *Lactobacillus,* and *Ruminococcus* were increased at all timepoints in post-AHFR-BLI subjects, *Finegoldia, Corynebacterium,* and *Prevotella* dominated throughout in the gut of the non-AHFR-BLI group (**Figure 2E**). Temporal analysis on differentially abundant taxa in both the URT and the gut revealed similar results (**Tables E4 and E5**).

Taken together, COVID-19 related AHFR-BLI induced long lasting effects on bacterial diversity in the URT, which was, at least in part, independent of antibiotic use or invasive ventilation during hospitalisation. Moreover, bacterial community composition in the URT and gut were distinct relative to COVID-19-related AHFR-BLI status.

### Post-COVID-19 lung function recovery is linked to a distinct URT and gut microbiota

Previous studies revealed that a restrictive ventilatory impairment with decreased TLC, FVC, and DLCO persisted for 12 months in critically ill COVID-19 patients(6). We assessed the association between longitudinal lung function recovery trajectories and microbiota profiles in a subset of our cohort who attended all three follow-up visits (**Table E6**). Recovery trajectories were established per subject for TLC, FVC, DLCO, FEV_1_, and FEV_1_/FVC (**Figure 3A** and **Figure E5**).

**Figure 3.**
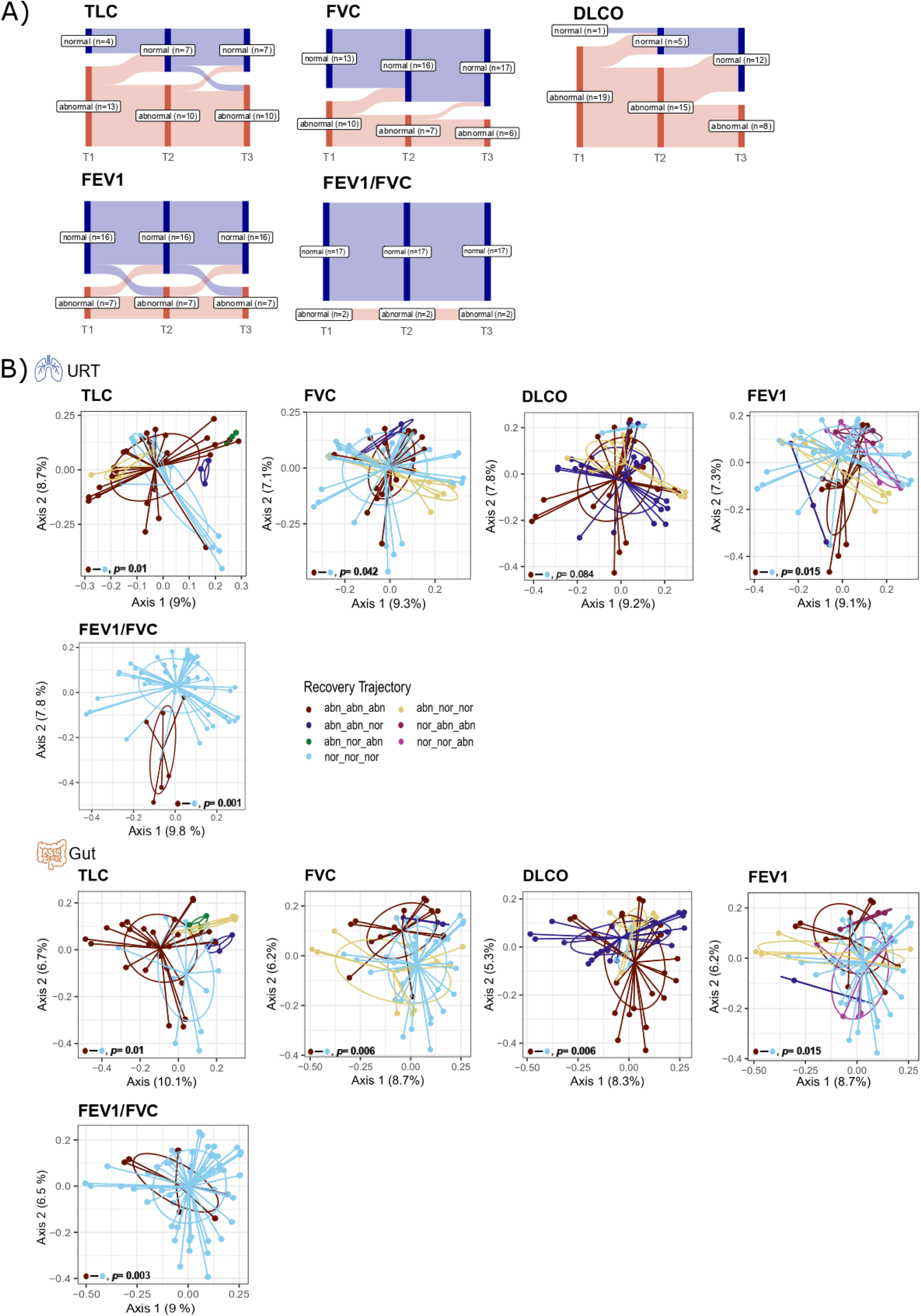
– Lung function recovery trajectories. (**A**) Sankey diagram representing lung function recovery trajectories over the study period, for each pulmonary function metric (TLC, FVC, DLCO, FEV_1,_ and FEV_1_/FVC). T1 = 3 -months, T2 = 6 -months, T3 = 12 -months post-hospitalisation. N = number of subjects. (**B**) Results of the principal coordinate analysis (PCoA) showing differences in beta-diversity (Bray-Curtis) between lung function recovery trajectories. Each dot represents one sample and each quadrant represents one pulmonary function metric. URT (top) and gut (bottom). Axis 1 and 2 indicate the proportion of variance. Comparisons were made using pairwise adonis, significance levels are presented in **Tables E7** and **E8**.

Bacterial community composition in both the URT and gut was significantly distinct relative to lung function recovery trajectory. In the URT, subjects having normal TLC, FVC, FEV_1_ and FEV_1_/FVC at all three follow-up visits (nor_nor_nor) could be distinguished from subjects having abnormal values at each visit (abn_abn_abn) (**Figure 3B**) (PERMANOVA, (TLC) *P* = 0.01, (FVC) *P* = 0.042, (FEV_1_) *P* = 0.015, (FEV_1_/FVC) *P* = 0.001). Similar findings were observed relative to the gut microbiota (PERMANOVA, (TLC) *P* = 0.01, (FVC), *P* = 0.006, (FEV_1_) *P* = 0.015, (DLCO) *P* = 0.006, (FEV_1_/FVC) *P* = 0.003) (**Figure 3B**, multiple comparisons in **Tables E7** and **E8**), while most subjects had normal values for FEV_1_/FVC ratio throughout the study period (17 out of 19 subjects)(**Figure 3B)**, suggesting that the presence of ventilatory impairment is mostly of restrictive pattern. In addition, the URT microbiota of subjects with a normal recovery trajectory was enriched in *Megasphera*, *Atopobium*, *Porphyromonas* and *Alloprevotella*, whilst subjects not recovering harboured more *Oribacterium* and *Bacteroides* (**Figure E6A**). Moreover, a normal recovery trajectory was linked to higher relative abundance of *Bacteroides*, *Streptococcus*, *Blautia*, *Faecalibacterium* and *Ruminococcus* in the gut, and subjects with abnormal recovery trajectories harboured more *Corynebacterium*, *Prevotella_9*, *Finegoldia* and *Agathobacter* (**Figure E6B**). Similarly, differential abundance analysis comparing normal and abnormal recovery trajectories highlighted the presence of URT *Streptococcus*, *Prevotella, Alloprevotella* and *Veillonella* in patients recovering, whereas *Streptococcus, Leptotrichia* and *Prevotella_7* were more present in patients never recovering to normal lung function. In the gut, taxa belonging to *Prevotella_9* strongly dominated in subjects with persisting pulmonary function impairment (**Tables E9** and **E10**). Altogether, this is the first evidence indicating the association between lung function recovery after severe COVID-19 with URT and gut microbiota composition.

### Lung function is correlated with shared bacterial taxa between URT and gut

ARDS is associated with intestinal barrier impairment, which can contribute to translocation of gut bacteria to the URT(21). To assess whether long-term pulmonary sequelae in post-AHFR-BLI subjects can be linked to the gut-lung crosstalk, we first determined similarities between the URT and gut microbiota. URT and gut ASVs were compared at nucleotide level, leading to the identification of 465 identical bacterial taxa between the two sampling sites. Proportionally, these common URT-gut taxa (i.e. shared taxa) accounted for up to 30% of the taxa present in the URT, and their amount did not differ relative to the time-point post-hospitalisation or AHFR-BLI status (**Figure 4A**). Among these common bacterial taxa, *Veillonella*, *Streptococcus* and *Rothia* were the top 3 most abundant genera. Interestingly, no differences were observed in the relative abundance of these 3 genera, or of *Bacteroides,* based on AHFR-BLI-status (**Figure 4B**). The presence of URT-gut shared taxa in the URT was negatively correlated with local URT α-diversity at T1 and T3 (Chao1, (T1) *P* = 0.018, (T2) *P* = 0.49, (T3) *P* = 0.025) (**Figure 4C**), suggesting that these common taxa disrupt the URT microbiota by reducing the local diversity.

**Figure 4.**
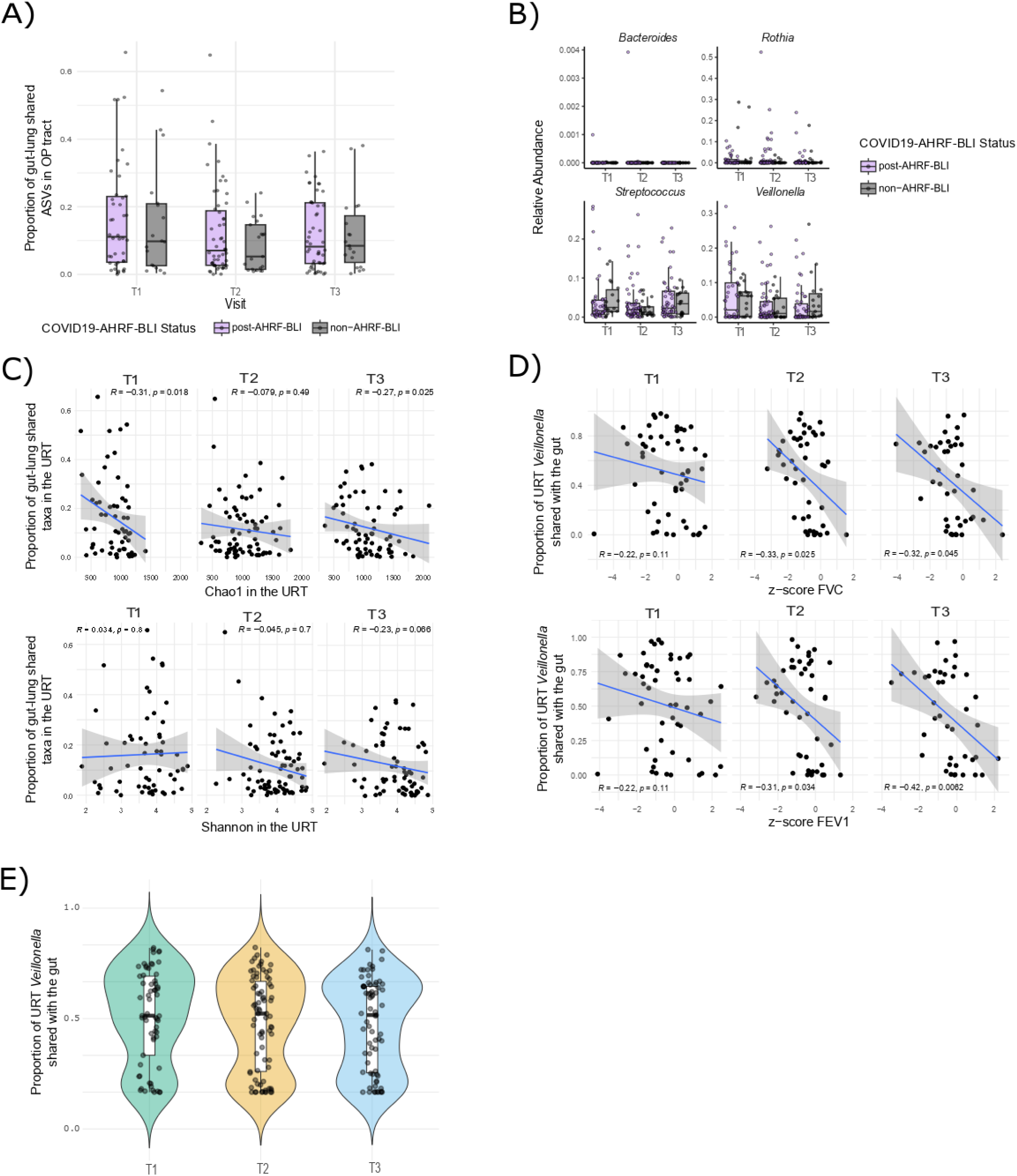
– Gut-lung axis. (**A**) Boxplot representing the proportion of URT-gut shared ASVs that are present in the URT at each follow-up visit, per AHRF-BLI-status. Each dot represents one sample. T1 = 3 -months, T2 = 6 -months, T3 = 12 -months post-hospitalization. Purple = post-AHRF-BLI, black = non-AHRF-BLI. Comparisons were made using Wilcoxon-test. (**B**) Relative abundance of *Bacteroides* and the top 3 genera among the URT-gut shared ASVs (*Veillonella*, *Streptococcus* and *Rothia*), per follow-up visit and per AHRF-BLI-status. Each dot represents one sample. T1 = 3 -months, T2 = 6 -months, T3 = 12 -months post-hospitalization. Purple = post-AHRF-BLI, black = non-AHRF-BLI. Comparisons were made using Wilcoxon-test. (**C**) Scatterplot representing the correlation between URT alpha diversity and the proportion of URT-gut shared ASVs in the URT at each follow-up visit. T1 = 3 -months, T2 = 6 -months, T3 = 12 -months post-hospitalization. Chao1 (top) and Shannon (bottom). Spearman was used to test for correlations. (**D**) Scatterplot representing the correlation between the proportion of URT *Veillonella* that is shared with the gut and lung function, per follow-up visit. T1 = 3 -months, T2 = 6 -months, T3 = 12 -months post-hospitalization. Z-score FVC (top) and z-score FEV_1_ (bottom). Spearman was used to test for correlations. (**E**) Violin -plot illustrating the proportion of *Veillonella* in the URT that is shared with the gut, per follow-up visit. Each dot represents one sample. T1 = 3 -months, T2 = 6 -months, T3 = 12 -months post-hospitalization. Comparisons were made using Kruskal-Wallis with Dunn’s post-hoc test and Bonferroni adjustment.

The relative abundance of URT *Veillonella* also found in the gut was negatively correlated with both FVC and FEV_1_ at T2 and T3 (FVC (T2) *P* = 0.025, (T3) *P* = 0.045, FEV_1_ (T2) *P* = 0.034, (T3) *P* = 0.0062) (**Figure 4D**), whereas no correlations were observed with TLC, DLCO, or FEV_1_/FVC at any timepoint. The abundance of URT-gut shared *Veillonella* was stable over the course of recovery (**Figure 4E**). The relationship between FVC or FEV_1_ and shared *Veillonella* independent of timepoint is non-linear, with a U-shaped association (**Figure E7A** and **B**). In a mixed-effects model with a random intercept for each patient, the abundance of URT–gut shared *Veillonella* was significantly associated with the non-linear component of FVC (*P* = 0.015), but not with FEV_1_ (**Table E11**). This finding indicates a non-linear relationship between shared *Veillonella* abundance and FVC, independent of potential confounders such as antibiotic or corticosteroid treatment during hospitalization.

Overall, these results suggest that the presence of shared *Veillonella* taxa between the URT and the gut is associated with lung function, indicating a potential key role of shared taxa between the airways and the gut in lung function recovery after severe COVID-19.

### Identification of URT mycobiota clusters with distinct bacterial composition

COVID-19 has been associated with secondary fungal infections, and little is known about the long-term influence of COVID-19 on airway fungal community composition. Investigation of the impact of severe COVID-19 on the URT mycobiota revealed that fungal diversity and load remained stable over the 12-month study period (**Figure E8A** and **B**).

Using unsupervised clustering, we identified 3 distinct mycobiota (ITS) clusters, where cluster 1 was dominated by *Saccharomyces*, cluster 2 by *Candida* and cluster 3 by *Cladosporium* (**Figure 5A and B**). While differences in composition between clusters were statistically significant (PERMANOVA *P* = 0.001), this may be partially attributable to the high variability in cluster 2 (PERMDISP *P* <0.001.) ITS load was significantly lower in cluster 3 compared to all others (**Figure E8C**), and cluster 2 was characterised by the lowest fungal α-diversity (**Figure 5C**). Although subject demographics as well as lung function did not differ between mycobiota clusters (**Table E12**), bacterial composition in cluster 2 was distinctly different from cluster 1 and 3 (**Figure 5D**, pairwise (adjusted) PERMANOVA, *P* = 0.003 for all pairs), with cluster 2 harbouring more *Rothia* and *Streptococcus*. In a mixed-effects model with a per-patient random intercept, we found that the increase in *Rothia* in cluster 3 was negatively correlated with FVC (**Table E13**). Additionally, *Neisseria* and *Veillonella* had statistically significant negative interactions with cluster 2. The potential confounding factors of antibiotic or corticosteroids were not significant, suggesting a possible link between *Rothia*, *Neisseria* and *Veillonella* with FVC. Only *Neisseria* and *Veillonella* maintained their significant interactions with ITS cluster 2 for FEV_1_ (**Table E14)**, and only *Veillonella* maintained this interaction with cluster 2 for TLC and DLCO (**Tables E15-E17**).

**Figure 5.**
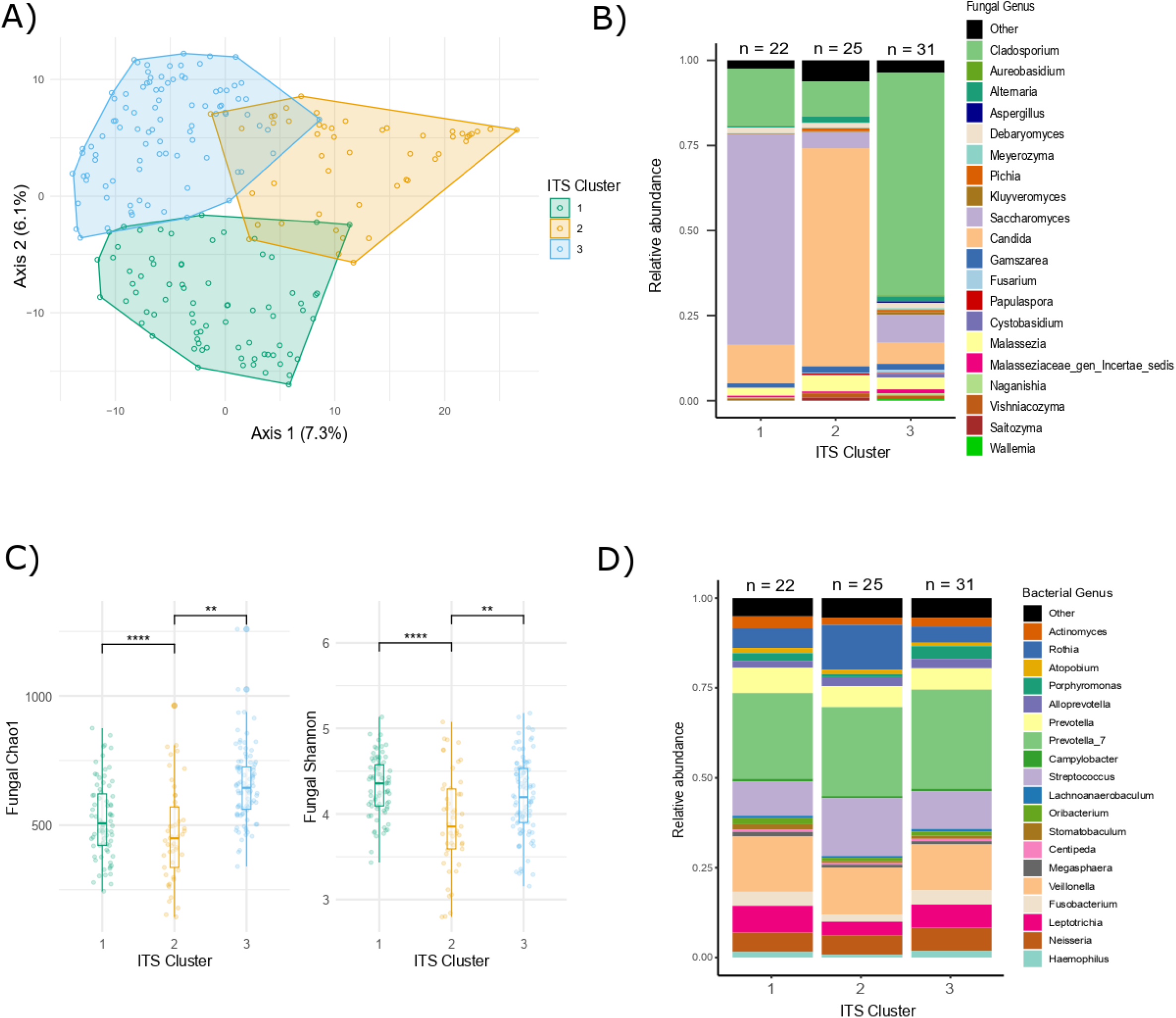
– URT Mycobiota clusters. (**A**) Results of the principal coordinate analysis (PCoA) showing differences in bacterial beta - diversity (Bray-Curtis) between ITS clusters. Each dot represents one sample. Axis 1 and 2 indicate the proportion of variance. Comparisons were made using adonis. **(B)** Relative abundance of the top 20 most abundant fungal genera per ITS cluster. The “other” category summarizes all remaining genera. N = number of subjects per group. (**C**) Boxplot representing alpha diversity per ITS cluster. Chao1 (left) and Shannon (right). Each dot represents one sample. Comparisons were made using Kruskal-Wallis with Dunn’s post-hoc test and Bonferroni adjustment. *P-value significance: *. < 0.05, **. ≤ 0.01, ***. ≤ 0.001.* (**D**) Relative abundance of the top 20 most abundant bacterial genera per ITS cluster. The “other” category summarizes all remaining genera. N = number of subjects per group.

Taken together, we identified three mycobiota clusters in our cohort, characterised by a distinct bacterial composition and correlation with lung function. The latter suggests the potential role of bacterial-fungal interactions in lung function recovery after severe COVID-19.

### Inter-kingdom interactions are distinct to each mycobiota cluster

Inter-kingdom interactions were assessed by constructing co-occurrence networks for each ITS cluster using the SPIEC-EASI algorithm. Visualization of cluster-specific vertices and their nearest neighbors revealed distinct bacterial–fungal associations (**Figure 6A**). Cluster 1 network was defined by two *Saccharomyces*-associated groups, cluster 2 by a single *Candida*-associated group, and cluster 3 by a broader pattern comprising *Gamszarea*-, *Malassezia*-, *Cladosporium-* and *Saccharomyces*-associated groups. Using a combined co-occurrence network including all samples, we determined the presence or absence of edges on a per-sample basis. Clusters 1 and 3 exhibited significantly higher numbers of fungi-fungi edges, with no significant differences observed for bacteria–bacteria or inter-kingdom edges (**Figure 6B** and **Table E18**). Fungal genera specific inter-kingdom edges (fungal-genera to bacteria-edges) were significantly higher for *Saccharomyces* in cluster 1, *Candida* in cluster 2, and *Cladosporium* in cluster 3 (**Table E19**).

**Figure 6.**
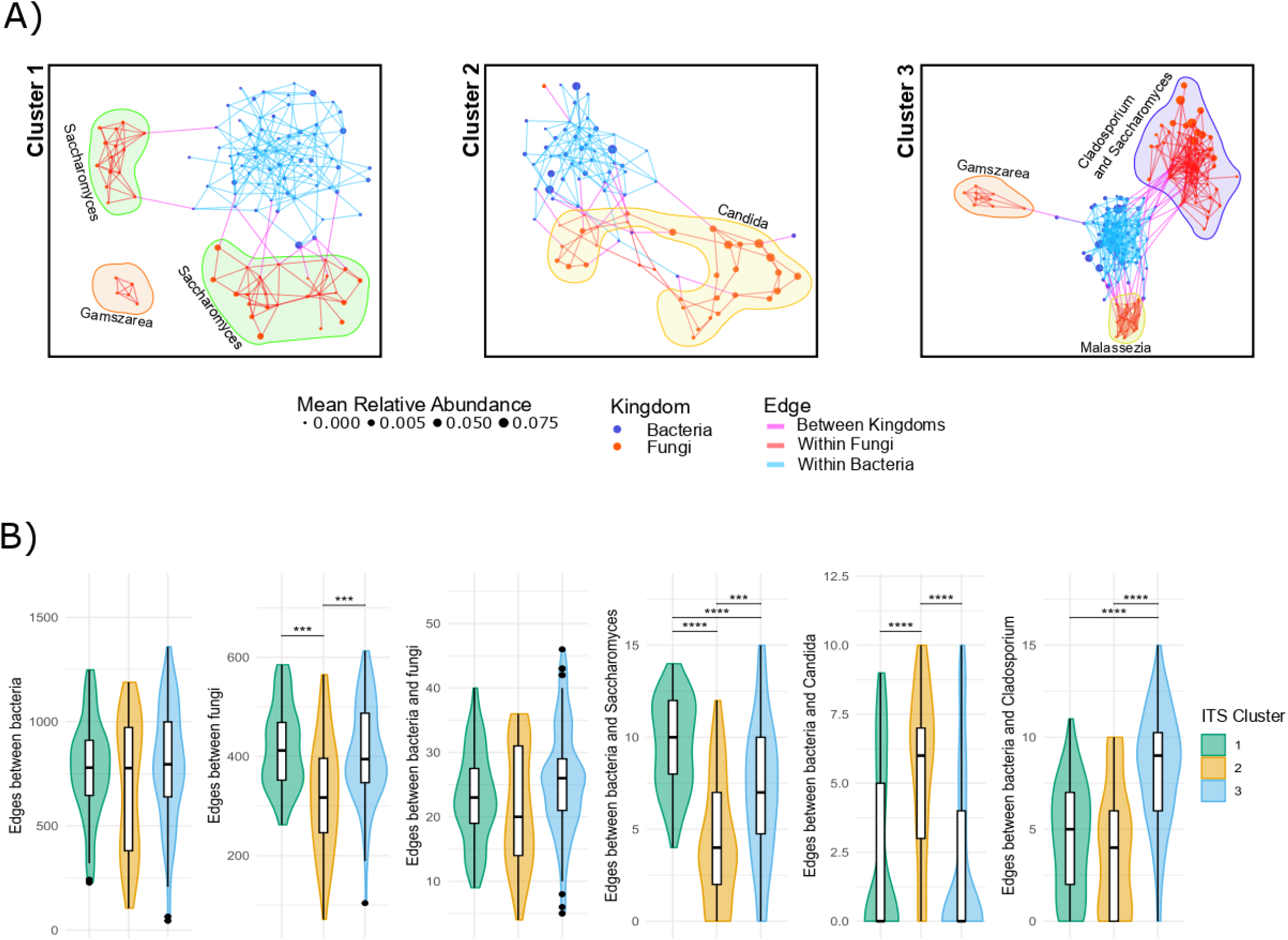
– URT Inter-kingdom analysis. **(A)** ITS clusters inter-kingdom graph connectivity. Sub-network graphs for ITS clusters. Graphs were generated with ASVs found in half all samples for a specific timepoint. The graphs were then further subsetted to unique vertices and their nearest neighbours. Point size indicates ASV average relative abundance, while kingdom is differentiated by color. Edges are colored by either within or between kingdoms. Labels are at genus level. (**B)** Graph intra- and inter-kingdom edge ITS cluster comparisons. Box and violin plots for edge numbers for each ITS cluster and edge type. Wilcoxon-test with Holm correction was used for comparisons (see **Table E16**). Means and standard deviations for each group are reported in **Table E16**. *P-value significance: *. < 0.05, **. ≤ 0.01, ***. ≤ 0.001*.

These results demonstrate that ITS clusters differ not only in community composition, but also in their inter-kingdom co-occurrence signatures.

## Discussion

Studies on long-term health outcomes after severe COVID-19 have reported persistent lung function abnormalities up to 1 year post-disease(4,6,38), yet the underlying mechanisms remain unclear. This study offers a comprehensive longitudinal analysis of the URT and gut microbiota following severe COVID-19, showing persistent, organ-specific dysbiosis associated with AHFR-BLI and lung function recovery.

In our cohort, URT *Veillonella* and *Alloprevotella* were enriched in individuals with improving lung function, corroborating previous findings in COPD(39). The presence of URT *Prevotella* and *Streptococcus* in both recovering and non-recovering patients highlights the heterogeneous role of these commensals, which can act as opportunistic pathogens in chronic respiratory disease(40–42). Additionally, *Blautia* and *Ruminococcus* - taxa described as potentially protective(43,44) - were more abundant in the gut of recovering subjects. Consistent with our results, post-COVID-19 patients with impaired lung function at 3 months showed reduced gut *Ruminococcus* and increased *Veillonella*(45). The observed loss of beneficial bacteria and outgrowth of potential pathobionts in non-recovering subjects suggest a key role of URT and gut microbiota in modulating lung function recovery after severe COVID-19, although the causative effect cannot be assessed in this study.

Several subjects in our cohort experienced AHFR-BLI, but did not fit the ARDS definition (2). Similary, ARDS has been linked to acute microbiota alterations and translocation of gut-associated bacteria (*Bacteroides spp*., *Lachnospiraceae*, *Enterobacteriaceae*) into the lungs(21,46). This study is the first to examine the long-term impact of COVID-19-related-AHFR-BLI on the URT and gut microbiota. AHFR-BLI was associated with sustained decreases in airway diversity, while the gut microbiota recovered more rapidly. Given the higher bacterial biomass in the gut, it is likely more resilient to respiratory viral infection. URT dysbiosis in COVID-19 patients preceding and exceeding that of the gut was previously described(47). Microbiota composition was distinct based on AHFR-BLI status, emphasizing AHFR-BLI’s long-term effects on both respiratory and gut microbiota. Unlike in acute COVID-19, we did not observe increased gut bacteria in the URT of post- AHFR-BLI subjects, but a negative correlation existed between shared gut-airway taxa—especially *Veillonella*—and lung function. These findings align with previous research showing increased gut *Veillonella* in COVID-19 survivors with impaired lung function at 3 months(45). The presence of oral *Veillonella* in COVID-19 subjects with ongoing symptoms might be linked to its pro-inflammatory role (48,49). Moreover, *Veillonella* in the gut and lung has been linked to idiopathic pulmonary fibrosis(50) and cystic fibrosis(51), highlighting the significance of the gut-lung axis in post-COVID-19 sequelae. Further research is needed to clarify the mechanistic role of *Veillonella* and other bacterial taxa in this setting.

Fungal communities have been implicated in COVID-19 pathogenesis(18,52). Analysis of URT fungal communities identified three distinct mycobiota clusters dominated by *Saccharomyces*, *Candida*, and *Cladosporium*, each associated with specific bacterial profiles. This suggests a fungal fingerprint among patient groups, characterized by a dominant fungus alongside a distinct microbiota. Such profiles may explain individual recovery trajectories after severe SARS-CoV-2 infection. Our mixed-effects model found a significant correlation between lung function and *Rothia* in subjects with *Cladosporium*-dominated URT, providing first evidence of bacterial-fungal interplay affecting lung function recovery. Previously, *Streptococcus* and *Rothia* have been linked to increased COPD exacerbation and emphysema risk(53). The presence of distinct airway fungi may partially explain bacterial behaviors, highlighting the essential role of airway mycobiota in unraveling respiratory tract host-bacterial-fungal interactions, which remain insufficiently explored.

Our study has several limitations. Conducted during the early COVID-19 pandemic, it was performed amid evolving and heterogeneous treatment guidelines. While medication may confound microbiota results, we demonstrated that our findings were not solely attributable to treatments. Similarly, we used the broader syndrome of „severe acute hypoxemic respiratory failure” to describe the most severely COVID-19 affected subjects. In fact, non-intubated and severe hypoxemic patients did not meet the latest Berlin definition of ARDS requiring a minimum PEEP of ≥ 5 cmH₂O, due to concerns regarding viral transmission from high-flow nasal oxygen (HFNO) in the early phase of the pandemic (3,54). This subgroup was therefore classified as having AHFR-BLI rather than ARDS (3,54), although they would have been treated with HFNO and thus classified as ARDS once risk for infectious spread was refuted.

Follow-up visits were based on individual clinical necessity; therefore, some study subjects did not have three time-points. Nonetheless, the distinct longitudinal microbiota profiles between recovered and non-recovered patients suggest microbiota involvement in long-term outcomes. The study’s design does not clarify causality, leaving it unclear whether microbiota influence recovery or are a risk factor for pulmonary sequelae. Lastly, valid comparison of microbiota studies remains complex due to variabilities in sampling site, sequencing methods, PCR conditions and the targeted 16S rRNA and ITS region(55). Despite these limitations, our results align with previous research, underscoring their clinical relevance.

In conclusion, our results reveal that persistent, compartment-specific microbiota alterations accompany COVID-19-related-AHFR-BLI, and govern pulmonary function recovery after severe COVID-19. Notably, common gut-airway bacteria such as *Veillonella* in the URT negatively correlate with lung function, highlighting the gut-lung axis’s role in lung function recovery after severe COVID-19. These findings offer novel insights into the URT and gut microenvironments’ contributions to long-term pulmonary sequelae, and open avenues for preventive and therapeutic strategies targeting severe SARS-CoV-2 infection, and potentially other aetiologies of viral lung infections.

## Data Availability

All data produced in the present study are available upon reasonable request to the authors

## Acknowledgements

We thank the patients who participated in the study, and Maryline Wiasemsky, Estelle Clément, Louis Mercier, Julie Pernot, Berra Erkosar, Alexandra Lenoir, Maura Prella, Catherine Beigelman-Aubry, and Brice Touilloux for contribution to patient inclusion, sample processing and data acquisition. 16S rRNA and ITS amplicon sequencing was performed at the Lausanne Genomic Technologies Facility, University of Lausanne, Switzerland (https://www.unil.ch/gtf/en/home.html).

## Authors’ contributions

*Conception and design*: EB, MFC, CVG, NDU. *Funding acquisition*: SVS, EB, MFC, CVG, NDU. *Acquisition of data*: SVS, LCS, EB, SG, AT, LP, MFC, CVG. *Analysis and interpretation of data:* SVS, LCS, EB, AT, LP, MFC, CVG and NDU. *Drafting or revising of manuscript:* SVS, LCS, CVG and NDU*. All authors critically revised and approved the final manuscript*.

## Funding sources

Bourse Solis (S.V.S), Lungenliga Bern (M.F.C.), Fondation Juchum and Fondation Placide Nicod (C.V.G) and the Ligue Pulmonaire Vaudoise (S.V.S and N.D.U.) contributed to the funding of the study but had no role in its design, conduct or analysis.

## Notes

### Competing Interest Statement

The authors have declared no competing interest.

### Funding Statement

This study was funded by: Bourse Solis (S.V.S), Lungenliga Bern (M.F.C.), Fondation Juchum and Fondation Placide Nicod (C.V.G) and the Ligue Pulmonaire Vaudoise (S.V.S and N.D.U.). All funders contributed to the funding of the study but had no role in its design, conduct or analysis.

### Author Declarations

The study was approved by the Swiss Ethics Committees on research involving humans (KEK 2020-00799, Identifier NCT04581135

