## Supplemental text and figures for "Respiratory and Gut Microbiota Correlate with Lung Function Recovery after Severe COVID-19"

### shared first author; * shared last author

#### Materials and Methods

##### *DNA isolation and quantification*

DNA was extracted from oropharyngeal and rectal swabs using the DNeasy UltraClean Microbial Kit (QIAGEN, Hilden, Germany), including blank DNA extraction controls (reagent controls). according to manufacturer's protocol with minor modifications. The protocol was modified by pre-incubation of the samples with 300 U Lyticase (Sigma- Aldrich, Germany) for 30 minutes at 37°C with gentle shaking (500 rpm) to enhance fungal DNA retrieval. The purified DNA was eluted in 40 µl of microbial DNA-free water (Qiagen).

Real-time quantitative PCR relying on previously published universal primers was used to determine the total copy number of the 16S rRNA gene(1) and the ITS region(2). We performed amplification using SsoAdvanced Universal SYBR Green Supermix (Bio-Rad, Hercules, CA, USA) on a Biorad CFX96 Real-Time detection machine with the following cycling parameters: initial 2 min denaturation at 98 °C, followed by 44 cycles of 5 s denaturation at 98 °C, and 60s annealing/elongation at 61.5 °C. Absolute quantification was performed based on a standard curve obtained with a purified amplicon product.

##### *16S rRNA amplicon sequencing*

Bacterial composition was assessed by Illumina MiSeq sequencing, using barcoded primers targeting the V1-V2 region. Amplification was performed with the Accuprime Taq DNA Polymerase High Fidelity kit (Invitrogen, Waltham, MA, USA). For the oropharyngeal swabs, duplicate PCR reactions of 20 µl consisted of 2 µl of 10× Accuprime buffer II, 0.44 µl of each 10 mM barcoded primer F-27 and R-338, 9.03 µl of ultrapure water, 0.09 µl of AccuPrime Taq DNA Polymerase and 8 µl of DNA template. The same steps were followed for the rectal swabs: duplicate PCR reactions of 30 µl consisted of 3 µl of 10× Accuprime buffer II, 0.67 µl of 10 µM barcoded primer F-27 and 6.7 µl of 1 µM R-338, 13.51 µl of ultrapure water, 0.13 µl of AccuPrime Taq DNA Polymerase and 6 µl of equimolar diluted DNA template.

Amplification was performed with the following cycling parameters: initial 3 min denaturation at 94 °C, followed by 40 cycles of 30 s denaturation at 94 °C, 30 s annealing at 56 °C and 90 s elongation at 72 °C, with a final extension at 72 °C for 5 min. We included no-template PCR reaction controls. Amplicons were quantified using a LabChip GX instrument with the DNA 1 K kit (Perkin Elmer, Waltham, MA, USA), pooled into equimolar amounts and purified using the AMPure XP bead cleaning system (Beckman Coulter, Brea, CA, USA). Libraries were diluted to 12 pM and spiked with 25% phiX before being loaded onto the Illumina MiSeq platform using pairwise chemistry, generating 250 × 2 read lengths (Lausanne Genomic Technologies Facility, University of Lausanne, Switzerland).

***Fungal ITS amplicon sequencing***

The ITS region from the extracted DNA was amplified by a 2-stage PCR. The first PCR was a multiplexed PCR, using 8 forward and 7 reverse primers(3,4). The locus specific sequences can be found in supplements **Table E1**. Amplification was performed using Accuprime Taq DNA Polymerase High Fidelity kit (Invitrogen, Waltham, MA, USA). The forward and reverse primers (10 µM) were respectively pooled with ultrapure water to a final concentration of 1 uM. Duplicate PCR reactions of 25 µl consisted of 5 µl forward primer pool (1 µM), 5 µl reverse primer pool (1 µM), 2.5 µl of 10× Accuprime buffer II, 6.4 µl of ultrapure water, 0.1 µl of AccuPrime Taq DNA Polymerase and 6 µl of DNA with the following cycling parameters: initial 3 min denaturation at 94 °C, followed by 40 cycles of 30 s denaturation at 94 °C, 30 s annealing at 55 °C and 60 s elongation at 68 °C, with a final extension at 68 °C for 5 min. We included no-template PCR reaction controls (n = 7). The resulting amplicons were subsequently purified (AMPure XP bead cleaning system (Beckman Coulter, Brea, CA, USA), quantified using a LabChip GX instrument with the DNA 1 K kit (Perkin Elmer, Waltham, MA, USA), and re-amplified through a second PCR which added barcodes and adaptor sequences. Amplicons were subsequently quantified using a LabChip GX instrument with the DNA 1 K kit (Perkin Elmer, Waltham, MA, USA), pooled into equimolar amounts and purified using the AMPure XP bead cleaning system (Beckman Coulter, Brea, CA, USA). Libraries were diluted to 12 pM and spiked with 25% phiX before being loaded onto the Illumina MiSeq platform using pairwise chemistry, generating 250 × 2 read lengths (Lausanne Genomic Technologies Facility, University of Lausanne, Switzerland).

*Bioinformatics and statistical analysis*

Samples with fewer than 10’000 reads 16S, or 2’000 reads ITS were removed. The SILVA (v138.1, 99% identity) and UNITE (v9.0) databases were used for taxonomic assignment of the 16S and ITS datasets respectively. Microbiome composition analysis was performed using the phyloseq package (v1.48.0)(5). Alpha diversity metrics Chao1 and Shannon were calculated using the microbiome package (v1.26.0)(6). Counts were transformed to relative abundance. The Bray-Curtis distance was calculated using the vegan package (v2.6.4) and visualized by principal coordinate analysis (PCoA) with the ggordiplots (v0.4.2) package. DESeq2 1.43.4 was used to evaluate the presence of differentially abundant taxa between groups.

#### Supplementary tables

**Table E1** Locus specific sequences (5’ – 3’) of the forward (fwd) and reverse (rev) Internal Transcriber Sequence (ITS) primers used for ITS amplicon sequencing.

| Primer | Sequence (5` - 3`) |
|---|---|
| ITS_fwd_1 | CTTGGTCATTTAGAGGAAGTAA |
| ITS_fwd_2 | CTCGGTCATTTAGAGGAAGTAA |
| ITS_fwd_3 | CTTGGTCATTTAGAGGAACTAA |
| ITS_fwd_4 | CCCGGTCATTTAGAGGAAGTAA |
| ITS_fwd_5 | CTAGGCTATTTAGAGGAAGTAA |
| ITS_fwd_6 | CTTAGTTATTTAGAGGAAGTAA |
| ITS_fwd_7 | CTACGTCATTTAGAGGAAGTAA |
| ITS_fwd_8 | CTTGGTCATTTAGAGGTCGTAA |
| ITS_rev_1 | GCTGCGTTCTTCATCGATGC |
| ITS_rev_2 | GCTGCGTTCTTCATCGATGG |
| ITS_rev_3 | GCTACGTTCTTCATCGATGC |
| ITS_rev_4 | GCTGCGTTCTTCATCGATGT |
| ITS_rev_5 | ACTGTGTTCTTCATCGATGT |
| ITS_rev_6 | GCTGCGTTCTTCATCGTTGC |
| ITS_rev_7 | GCGTTCTTCATCGATGC |

**Table E2** Demographics of the study cohort per COVID-19 -related AHFR-BLI status. N = number of subjects per group.

| **Characteristic** | **Overall**, N = 83[1] | **COVID-19 non- AHFR-BLI**, N = 22[1] | **COVID-19 post- AHFR-BLI**, N = 61[1] | ***P*-value** |
|---|---|---|---|---|
| **Age** (yr) | 62 (57, 71) | 61 (57, 67) | 62 (56, 71) | 0.82[2] |
| **BMI** (kg/m$^2$) | 29.0 (26.0, 32.3) | 27.5 (25.5, 29.3) | 29.7 (26.5, 33.2) | 0.043[2] |
| **Gender** | | | | 0.30[3] |
| Female | 22 (27%) | 4 (18%) | 18 (30%) | |
| Male | 61 (73%) | 18 (82%) | 43 (70%) | |
| **Intubation** | 49 (60%) | 1 (4.8%) | 48 (79%) | <0.001[3] |
| **Antibiotics** | 61 (73%) | 9 (41%) | 52 (85%) | <0.001[3] |
| **Corticosteroids** | 39 (48%) | 18 (82%) | 21 (36%) | <0.001[3] |
| **Anti-IL6Ri** (Tocilizumab) | 31 (37%) | 5 (23%) | 26 (43%) | 0.10[3] |
| **Asthma** | 11 (13%) | 4 (18%) | 7 (11%) | 0.47[4] |
| **COPD** | 2 (2.4%) | 1 (4.5%) | 1 (1.6%) | 0.46[4] |
| **Lung Cancer** | 2 (2.4%) | 1 (4.5%) | 1 (1.6%) | 0.46[4] |
| **Smoking Status** | | | | 0.64[3] |
| Ever | 32 (41%) | 8 (36%) | 24 (42%) | |
| Never | 47 (59%) | 14 (64%) | 33 (58%) | |

[1]Median (IQR) or n (%);[2]Wilcoxon rank sum test; [3]Pearson's Chi-squared test; [4]Fisher's exact test

**Table E3** Per timepoint permanova (999 permutations) of Bray-Curtis beta diversity for Intubation or Antibiotics in the upper respiratory tract (URT) and gut.

| Timepoint | Df | SumOfSqs | R2 | F | Pr(>F) |
|---|---|---|---|---|---|
| **Intubation (URT)** | | | | | |
| T1 | 1 | 0,3696 | 0,0197 | 1,2275 | 0,072 |
| T2 | 1 | 0,348 | 0,0144 | 1,1548 | 0,126 |
| T3 | 1 | 0,3402 | 0,0155 | 1,1358 | 0,173 |
| **Intubation (Gut)** | | | | | |
| T1 | 1 | 0,5187 | 0,0237 | 1,3833 | 0,013 |
| T2 | 1 | 0,4879 | 0,0168 | 1,2658 | 0,029 |
| T3 | 1 | 0,4375 | 0,0172 | 1,1567 | 0,093 |
| **Antibiotics (URT)** | | | | | |
| T1 | 1 | 0,4261 | 0,0224 | 1,4208 | 0,014 |
| T2 | 1 | 0,3278 | 0,0134 | 1,0887 | 0,229 |
| T3 | 1 | 0,2955 | 0,0133 | 0,9871 | 0,445 |
| **Antibiotics (Gut)** | | | | | |
| T1 | 1 | 0,4913 | 0,022 | 1,3055 | 0,028 |
| T2 | 1 | 0,6279 | 0,0214 | 1,6365 | 0,003 |
| T3 | 1 | 0,4189 | 0,0162 | 1,1048 | 0,187 |

**Table E4 DESEq2 results, ASVs – URT – AHFR-BLI (excel)**

**Table E5 DESEq2 results, ASVs – Gut – AHFR-BLI (excel)**

**Table E6** Demographics table of subjects with available lung function metric (TLC, FVC, FEV1 and DLCO) and having attended all three follow-up visits. N = number of subjects per group.

| **Characteristic** | *TLC* **N = 17**[1] | *FVC* **N = 23**[1] | *DLCO* **N = 20**[1] | *FEV$_1$* **N = 23**[1] |
|---|---|---|---|---|
| **Age** | 62 (59, 71) | 61 (52, 69) | 61 (57, 67) | 61 (52, 69) |
| **BMI** | 28.4 (25.6, 32.3) | 29.4 (26.2, 32.8) | 30.3 (27.8, 33.4) | 29.4 (26.2, 32.8) |
| **Gender** | | | | |
| Female | 2 (12%) | 4 (17%) | 5 (25%) | 4 (17%) |
| Male | 15 (88%) | 19 (83%) | 15 (75%) | 19 (83%) |
| **Intubation** | 10 (59%) | 14 (61%) | 12 (60%) | 14 (61%) |
| **COVID-19 AHFR-BLI** | 11 (65%) | 16 (70%) | 15 (75%) | 16 (70%) |
| **Antibiotics** | 12 (71%) | 17 (74%) | 13 (65%) | 17 (74%) |
| **Corticosteroids** | 11 (65%) | 15 (65%) | 12 (60%) | 15 (65%) |
| **Anti-IL6Ri (Tocilizumab)** | 6 (35%) | 7 (30%) | 6 (30%) | 7 (30%) |
| **Asthma** | 1 (5.9%) | 2 (8.7%) | 2 (10%) | 2 (8.7%) |
| **COPD** | 2 (12%) | 2 (8.7%) | 2 (10%) | 2 (8.7%) |
| **Lung Cancer** | 2 (12%) | 2 (8.7%) | 2 (10%) | 2 (8.7%) |
| **Smoking Status** | | | | |
| Ever | 9 (56%) | 10 (45%) | 9 (47%) | 10 (45%) |
| Never | 7 (44%) | 12 (55%) | 10 (53%) | 12 (55%) |

[1]Median (IQR) or n (%)

**Table E7** Significance levels of differences in URT beta -diversity (Bray-Curtis) between lung function recovery trajectories (T1_T2_T3, i.e. 3-, 6- and 12-months after hospitalisation). Comparisons were made using pairwise adonis. nor = normal, abn = abnormal lung function metric.

| Comparison | Sums of squares | F-Model | R2 | *P*-value | *P*-value adjusted | Sign. |
|---|---|---|---|---|---|---|
| ***TLC*** | | | | | | |
| nor_nor_nor **vs** abn_abn_abn | 0.634924953 | 1.984996534 | 0.05091693 | 0.001 | 0.01 | * |
| nor_nor_nor **vs** abn_nor_nor | 0.660834428 | 2.279143457 | 0.12468546 | 0.006 | 0.06 | |
| nor_nor_nor **vs** abn_abn_nor | 0.649898933 | 2.227288024 | 0.14626951 | 0.016 | 0.16 | |
| nor_nor_nor **vs** abn_nor_abn | 0.794511705 | 2.761783102 | 0.17522022 | 0.007 | 0.07 | |
| abn_abn_abn **vs** abn_nor_nor | 0.563263839 | 1.875666488 | 0.05705334 | 0.004 | 0.04 | . |
| abn_abn_abn **vs** abn_abn_nor | 0.727960963 | 2.408358057 | 0.07920053 | 0.001 | 0.01 | * |
| abn_abn_abn **vs** abn_nor_abn | 0.780099387 | 2.597241982 | 0.08488484 | 0.001 | 0.01 | * |
| abn_nor_nor **vs** abn_abn_nor | 0.660105212 | 3.645012368 | 0.34241504 | 0.012 | 0.12 | |
| abn_nor_nor **vs** abn_nor_abn | 0.777277993 | 4.480818532 | 0.39028738 | 0.012 | 0.12 | |
| abn_abn_nor **vs** abn_nor_abn | 0.613787227 | 6.665094772 | 0.62494473 | 0.1 | 1 | |
| ***FVC*** | | | | | | |
| nor_nor_nor **vs** abn_abn_nor | 0.553425454 | 1.759420912 | 0.04213231 | 0.02 | 0.12 | |
| nor_nor_nor **vs** abn_abn_abn | 0.522648382 | 1.663928846 | 0.02936487 | 0.007 | 0.042 | . |
| nor_nor_nor **vs** abn_nor_nor | 0.645011975 | 2.029073254 | 0.04224677 | 0.001 | 0.006 | * |
| abn_abn_nor **vs** abn_abn_abn | 0.531467855 | 1.894788005 | 0.09068233 | 0.013 | 0.078 | |
| abn_abn_nor **vs** abn_nor_nor | 0.629483746 | 2.352126277 | 0.19042278 | 0.019 | 0.114 | |
| abn_abn_abn **vs** abn_nor_nor | 0.569404533 | 1.931489857 | 0.07171864 | 0.008 | 0.048 | . |
| ***$FEV_1$*** | | | | | | |
| abn_abn_abn **vs** nor_nor_nor | 0.72190062 | 2.354249885 | 0.048687548 | 0.001 | 0.015 | . |
| abn_abn_abn **vs** abn_abn_nor | 0.570954219 | 1.778808048 | 0.120362078 | 0.022 | 0.33 | |
| abn_abn_abn **vs** nor_nor_abn | 0.727737371 | 2.518050216 | 0.13597815 | 0.001 | 0.015 | . |
| abn_abn_abn **vs** nor_abn_abn | 0.559385714 | 2.047186792 | 0.136051132 | 0.012 | 0.18 | |
| abn_abn_abn **vs** abn_nor_nor | 0.518214533 | 1.781962211 | 0.100211787 | 0.021 | 0.315 | |
| nor_nor_nor **vs** abn_abn_nor | 0.526660405 | 1.679497738 | 0.043420878 | 0.014 | 0.21 | |
| nor_nor_nor **vs** nor_nor_abn | 0.661575893 | 2.19537649 | 0.05202884 | 0.001 | 0.015 | . |
| nor_nor_nor **vs** nor_abn_abn | 0.507228435 | 1.708920441 | 0.044147975 | 0.008 | 0.12 | |
| nor_nor_nor **vs** abn_nor_nor | 0.529914804 | 1.754273581 | 0.042014228 | 0.015 | 0.225 | |
| abn_abn_nor **vs** nor_nor_abn | 0.589613938 | 1.94559218 | 0.217491714 | 0.049 | 0.735 | |
| abn_abn_nor **vs** nor_abn_abn | 0.620907334 | 2.366659132 | 0.371727005 | 0.1 | 1 | |
| abn_abn_nor **vs** abn_nor_nor | 0.532111293 | 1.732290722 | 0.198377582 | 0.03 | 0.45 | |
| nor_nor_abn **vs** nor_abn_abn | 0.558979719 | 2.607056139 | 0.271368888 | 0.027 | 0.405 | |
| nor_nor_abn **vs** abn_nor_nor | 0.628049402 | 2.414040458 | 0.194460495 | 0.013 | 0.195 | |
| nor_abn_abn **vs** abn_nor_nor | 0.505253196 | 2.312041289 | 0.24828512 | 0.043 | 0.645 | |
| ***DLCO*** | | | | | | |
| abn_abn_abn **vs** abn_nor_nor | 0.626806658 | 2.097895 | 0.058117 | 0.002 | 0.012 | . |
| abn_abn_abn **vs** abn_abn_nor | 0.56266258 | 1.780331 | 0.039757 | 0.004 | 0.024 | . |
| abn_abn_abn **vs** nor_nor_nor | 0.582787 | 1.9179 | 0.07125 | 0.014 | 0.084 | |
| abn_nor_nor **vs** abn_abn_nor | 0.586519443 | 2.005529 | 0.060763 | 0.002 | 0.012 | . |
| abn_nor_nor **vs** nor_nor_nor | 0.545205043 | 2.306589 | 0.150693 | 0.013 | 0.078 | |
| abn_abn_nor **vs** nor_nor_nor | 0.587484066 | 1.987126 | 0.082841 | 0.007 | 0.042 | . |

**Table E8** Significance levels of differences in gut beta -diversity (Bray-Curtis) between lung function recovery trajectories (T1_T2_T3, i.e. 3-, 6- and 12-months after hospitalisation). Comparisons were made using pairwise adonis. nor = normal, abn = abnormal lung function metric.

| Comparison | Sums of squares | F-Model | R2 | *P*-value | *P*-value adjusted | Sign. |
|---|---|---|---|---|---|---|
| ***TLC*** | | | | | | |
| nor_nor_nor vs abn_abn_abn | 0.77873993 | 2.116772861 | 0.055533895 | 0.001 | 0.01 | * |
| nor_nor_nor vs abn_nor_nor | 0.785905376 | 2.431019183 | 0.13189825 | 0.001 | 0.01 | * |
| nor_nor_nor vs abn_abn_nor | 0.780810611 | 2.345958847 | 0.152871441 | 0.011 | 0.11 | |
| nor_nor_nor vs abn_nor_abn | 0.789918913 | 2.412049651 | 0.156504145 | 0.005 | 0.05 | . |
| abn_abn_abn vs abn_nor_nor | 0.892276476 | 2.577036634 | 0.079105926 | 0.001 | 0.01 | * |
| abn_abn_abn vs abn_abn_nor | 0.891447004 | 2.5225558 | 0.085445035 | 0.001 | 0.01 | * |
| abn_abn_abn vs abn_nor_abn | 0.829140625 | 2.36345224 | 0.08048959 | 0.003 | 0.03 | . |
| abn_nor_nor vs abn_abn_nor | 0.775744671 | 3.693995068 | 0.345427041 | 0.018 | 0.18 | |
| abn_nor_nor vs abn_nor_abn | 0.789794381 | 3.947435212 | 0.360580824 | 0.006 | 0.06 | |
| abn_abn_nor vs abn_nor_abn | 0.7297433 | 5.260859021 | 0.568074626 | 0.1 | 1 | |
| ***FVC*** | | | | | | |
| nor_nor_nor vs abn_abn_nor | 0.824476935 | 2.357587723 | 0.061463399 | 0.002 | 0.012 | . |
| nor_nor_nor vs abn_abn_abn | 0.784500656 | 2.173150796 | 0.044193849 | 0.001 | 0.006 | * |
| nor_nor_nor vs abn_nor_nor | 0.808477932 | 2.242587862 | 0.050688442 | 0.001 | 0.006 | * |
| abn_abn_nor vs abn_abn_abn | 0.805557694 | 2.508595632 | 0.143277947 | 0.003 | 0.018 | . |
| abn_abn_nor vs abn_nor_nor | 0.873634117 | 2.920485917 | 0.226035301 | 0.019 | 0.114 | |
| abn_abn_abn vs abn_nor_nor | 0.793262631 | 2.260738114 | 0.09719116 | 0.002 | 0.012 | . |
| ***$FEV_1$*** | | | | | | |
| nor_nor_nor vs abn_abn_nor | 0.535657851 | 1.48726234 | 0.040761138 | 0.047 | 0.705 | |
| nor_nor_nor vs abn_nor_nor | 0.826272758 | 2.309575118 | 0.055908954 | 0.001 | 0.015 | . |
| nor_nor_nor vs abn_abn_abn | 0.806080906 | 2.262207899 | 0.051109242 | 0.001 | 0.015 | . |
| nor_nor_nor vs nor_nor_abn | 0.793021539 | 2.240713205 | 0.054332552 | 0.001 | 0.015 | . |
| nor_nor_nor vs nor_abn_abn | 0.774145802 | 2.200020887 | 0.057592138 | 0.002 | 0.03 | . |
| abn_abn_nor vs abn_nor_nor | 0.576204536 | 1.832555605 | 0.233966498 | 0.132 | 1 | |
| abn_abn_nor vs abn_abn_abn | 0.594345334 | 1.844787998 | 0.170108258 | 0.055 | 0.825 | |
| abn_abn_nor vs nor_nor_abn | 0.573031088 | 1.979824858 | 0.248103798 | 0.076 | 1 | |
| abn_abn_nor vs nor_abn_abn | 0.637760333 | 3.180030822 | 0.514565528 | 0.1 | 1 | |
| abn_nor_nor vs abn_abn_abn | 0.791078496 | 2.421777361 | 0.157036203 | 0.008 | 0.12 | |
| abn_nor_nor vs nor_nor_abn | 0.811131763 | 2.630547602 | 0.20826869 | 0.009 | 0.135 | |
| abn_nor_nor vs nor_abn_abn | 0.818160125 | 2.939166733 | 0.295715608 | 0.032 | 0.48 | |
| abn_abn_abn vs nor_nor_abn | 0.841581045 | 2.6706898 | 0.170425797 | 0.002 | 0.03 | . |
| abn_abn_abn vs nor_abn_abn | 0.755782609 | 2.551965471 | 0.203312021 | 0.01 | 0.15 | |
| nor_nor_abn vs nor_abn_abn | 0.760963613 | 2.961600787 | 0.297301694 | 0.022 | 0.33 | |
| ***DLCO*** | | | | | | |
| abn_abn_abn vs abn_nor_nor | 0.845890499 | 2.457210936 | 0.067399861 | 0.001 | 0.006 | * |
| abn_abn_abn vs abn_abn_nor | 0.885585457 | 2.423159037 | 0.053346335 | 0.001 | 0.006 | * |
| abn_abn_abn vs nor_nor_nor | 0.865669224 | 2.504885921 | 0.09107058 | 0.001 | 0.006 | * |
| abn_nor_nor vs abn_abn_nor | 0.807438625 | 2.306936359 | 0.069262941 | 0.002 | 0.012 | . |
| abn_nor_nor vs nor_nor_nor | 0.960097125 | 3.306418874 | 0.202767934 | 0.001 | 0.006 | * |
| abn_abn_nor vs nor_nor_nor | 0.924038819 | 2.611112725 | 0.106094867 | 0.002 | 0.012 | . |

**Table E9 DESEq2 results, ASVs – URT – lung function (excel)**

**Table E10 DESEq2 results, ASVs – Gut – lung function (excel)**

**Table E11** Fixed effects estimates from a mixed-effects model of FVC and FEV1 z-score by shared *Veillonella* and by medication administration during hospitalisation.

| Independent Variable | Coefficient (95% CI) | Std. Error | Statistic | df | *P*-value |
|---|---|---|---|---|---|
| **FVC** | | | | | |
| (Intercept) | -0.888 (-1.637 to -0.139) | 0.370 | -2.397 | 39.042 | 0.021 |
| polyn. shared *Veillonella* order:1 | -1.445 (-3.083 to 0.194) | 0.823 | -1.756 | 77.927 | 0.083 |
| polyn. shared *Veillonella* order:2 | 1.782 (0.353 to 3.211) | 0.717 | 2.486 | 72.206 | 0.015 |
| Antibiotics (Yes) | 0.116 (-0.651 to 0.883) | 0.379 | 0.306 | 38.290 | 0.761 |
| Corticosteroids (Yes) | -0.267 (-0.919 to 0.384) | 0.322 | -0.830 | 38.781 | 0.412 |
| Inhaled Corticosteroids (Yes) | 0.094 (-2.11 to 2.298) | 1.091 | 0.086 | 40.621 | 0.932 |
| **$FEV_1$** | | | | | |
| (Intercept) | -0.739 (-1.503 to 0.025) | 0.378 | -1.956 | 39.736 | 0.057 |
| polyn. shared *Veillonella* order:1 | -1.146 (-2.684 to 0.391) | 0.772 | -1.485 | 75.558 | 0.142 |
| polyn. shared *Veillonella* order:2 | 1.165 (-0.171 to 2.501) | 0.670 | 1.739 | 70.769 | 0.086 |
| Antibiotics (Yes) | 0.249 (-0.534 to 1.032) | 0.387 | 0.644 | 39.096 | 0.524 |
| Corticosteroids (Yes) | -0.152 (-0.816 to 0.513) | 0.329 | -0.461 | 39.518 | 0.647 |
| Inhaled Corticosteroids (Yes) | -0.567 (-2.811 to 1.676) | 1.111 | -0.511 | 41.045 | 0.612 |

**Table E12** Demographics table per ITS cluster. N = number of subjects per group.

| Characteristic | ITS cluster 1, N = 22[1] | ITS cluster 2, N = 25[1] | ITS cluster 3, N = 31[1] | *P*-value | q-value[2] |
|---|---|---|---|---|---|
| **Age** (yr) | 62 (57, 70) | 65 (58, 72) | 60 (55, 69) | 0.40[3] | >0.99 |
| **BMI** (kg/m$^2$) | 29.3 (25.9, 32.8) | 29.4 (25.8, 32.1) | 28.4 (26.1, 31.5) | 0.86[3] | >0.99 |
| **Gender** | | | | 0.26[4] | >0.99 |
| Female | 8 (36%) | 4 (16%) | 7 (23%) | | |
| Male | 14 (64%) | 21 (84%) | 24 (77%) | | |
| **Intubation** | 12 (55%) | 18 (75%) | 18 (58%) | 0.29[4] | >0.99 |
| **COVID-19 AHFR-BLI** | | | | 0.44[4] | >0.99 |
| COVID-19 non-AHFR-BLI | 6 (27%) | 5 (20%) | 11 (35%) | | |
| COVID-19 post-AHFR-BLI | 16 (73%) | 20 (80%) | 20 (65%) | | |
| **Antibiotics** | 18 (82%) | 16 (64%) | 23 (74%) | 0.38[4] | >0.99 |
| **Anti-IL6Ri** | 7 (32%) | 10 (40%) | 13 (42%) | 0.74[4] | >0.99 |
| **Corticosteroids** | 12 (55%) | 7 (30%) | 19 (61%) | 0.071[4] | 0.85 |
| **Asthma** | 2 (9.1%) | 4 (16%) | 4 (13%) | 0.91[5] | >0.99 |
| **COPD** | 0 (0%) | 2 (8.0%) | 0 (0%) | 0.18[5] | >0.99 |
| **Lung Cancer** | 0 (0%) | 2 (8.0%) | 0 (0%) | 0.18[5] | >0.99 |
| **Smoking Status** | | | | 0.49[4] | >0.99 |
| Ever | 7 (33%) | 11 (48%) | 10 (33%) | | |
| Never | 14 (67%) | 12 (52%) | 20 (67%) | | |
| **TLC (z-score)** | -1.38 (-1.95, -0.56) | -1.80 (-2.90, -0.75) | -1.75 (-2.39, -0.61) | 0.50[3] | >0.99 |
| **FVC (z-score)** | -0.77 (-1.55, 0.21) | -0.91 (-2.12, 0.29) | -0.52 (-1.59, 0.12) | 0.67[3] | >0.99 |
| **$FEV_1$ (z-score)** | -0.20 (-0.85, 0.75) | -0.94 (-1.62, -0.36) | -0.44 (-1.11, 0.29) | 0.11[3] | >0.99 |
| **DLCO (z-score)** | -1.71 (-2.31, -1.23) | -1.90 (-2.87, -1.15) | -1.81 (-2.60, -1.25) | 0.85[3] | >0.99 |

| Characteristic | ITS cluster 1, N = 22[1] | ITS cluster 2, N = 25[1] | ITS cluster 3, N = 31[1] | *P*-value | q-value[2] |
| --- | --- | --- | --- | --- | --- |

[1]Median (IQR) or n (%); [2]Bonferroni correction for multiple testing; [3]Kruskal-Wallis rank sum test; [4]Pearson's Chi-squared test; [5]Fisher's exact test

**Table E13** Fixed effects estimates from a mixed-effects model of FVC z-score, including ITS cluster, microbiota genera, and medication administration during hospitalisation. Interactions between independent variables are denoted by ':'.

| **Independent Variable** | **Coefficient (95% CI)** | **Std. Error** | **Statistic** | **df** | ***P*-value** |
|---|---|---|---|---|---|
| (Intercept) | -0.451 (-1.262 to 0.36) | 0.403 | -1.118 | 47.802 | 0.269 |
| polyn. shared *Veillonella* order:1 | -1.061 (-2.471 to 0.349) | 0.706 | -1.504 | 63.813 | 0.138 |
| polyn. shared *Veillonella* order:2 | 1.627 (0.376 to 2.879) | 0.625 | 2.602 | 59.810 | 0.012 |
| Antibiotics (Yes) | 0.061 (-0.713 to 0.834) | 0.382 | 0.159 | 37.222 | 0.875 |
| Corticosteroids (Yes) | -0.261 (-0.925 to 0.403) | 0.328 | -0.796 | 39.177 | 0.431 |
| Inhaled Corticosteroids (Yes) | -0.348 (-2.566 to 1.869) | 1.096 | -0.318 | 38.956 | 0.752 |
| ITS Cluster 1:*Rothia* | -0.886 (-3.096 to 1.323) | 1.110 | -0.798 | 79.288 | 0.427 |
| ITS Cluster 2:*Rothia* | -0.08 (-2.149 to 1.99) | 1.040 | -0.077 | 79.609 | 0.939 |
| ITS Cluster 3:*Rothia* | -7.507 (-14.014 to -1) | 3.258 | -2.304 | 64.557 | 0.024 |
| ITS Cluster 1:*Neisseria* | -0.888 (-2.86 to 1.085) | 0.988 | -0.898 | 67.495 | 0.372 |
| ITS Cluster 2:*Neisseria* | -5.59 (-7.809 to -3.371) | 1.112 | -5.025 | 69.649 | 0.000 |
| ITS Cluster 3:*Neisseria* | -1.151 (-3.014 to 0.712) | 0.932 | -1.235 | 62.127 | 0.221 |
| ITS Cluster 1:*Veillonella* | -0.525 (-2.443 to 1.393) | 0.961 | -0.546 | 69.552 | 0.587 |
| ITS Cluster 2:*Veillonella* | -3.329 (-5.535 to -1.123) | 1.106 | -3.009 | 71.582 | 0.004 |
| ITS Cluster 3:*Veillonella* | 0.271 (-2.051 to 2.593) | 1.164 | 0.233 | 68.056 | 0.817 |

**Table E14** Fixed effects estimates from a mixed-effects model of FEV1 z-score, including ITS cluster, microbiota genera, and medication administration during hospitalisation. Interactions between independent variables are denoted by ':'.

| **Independent Variable** | **Coefficient (95% CI)** | **Std. Error** | **Statistic** | **df** | ***P*-value** |
|---|---|---|---|---|---|
| (Intercept) | -0.306 (-1.135 to 0.523) | 0.412 | -0.742 | 48.245 | 0.462 |
| polyn. shared *Veillonella* order:1 | -0.713 (-2.06 to 0.634) | 0.674 | -1.058 | 62.935 | 0.294 |
| polyn. shared *Veillonella* order:2 | 1.053 (-0.138 to 2.245) | 0.596 | 1.769 | 59.507 | 0.082 |
| Antibiotics (Yes) | 0.222 (-0.576 to 1.02) | 0.394 | 0.563 | 38.574 | 0.576 |
| Corticosteroids (Yes) | -0.119 (-0.803 to 0.564) | 0.338 | -0.352 | 40.322 | 0.726 |
| Inhaled Corticosteroids (Yes) | -1.015 (-3.299 to 1.269) | 1.130 | -0.898 | 40.090 | 0.375 |
| ITS Cluster 1:*Rothia* | -1.809 (-3.942 to 0.324) | 1.071 | -1.689 | 76.404 | 0.095 |
| ITS Cluster 2:*Rothia* | -1.092 (-3.09 to 0.907) | 1.004 | -1.088 | 76.838 | 0.280 |
| ITS Cluster 3:*Rothia* | -5.845 (-12.063 to 0.373) | 3.112 | -1.878 | 63.768 | 0.065 |
| ITS Cluster 1:*Neisseria* | -1.178 (-3.067 to 0.711) | 0.946 | -1.245 | 66.082 | 0.218 |
| ITS Cluster 2:*Neisseria* | -4.774 (-6.902 to -2.646) | 1.066 | -4.477 | 68.010 | 0.000 |
| ITS Cluster 3:*Neisseria* | -1.544 (-3.321 to 0.233) | 0.889 | -1.737 | 61.525 | 0.087 |
| ITS Cluster 1:*Veillonella* | -0.068 (-1.907 to 1.771) | 0.922 | -0.074 | 67.778 | 0.942 |
| ITS Cluster 2:*Veillonella* | -3.266 (-5.384 to -1.148) | 1.062 | -3.076 | 69.859 | 0.003 |
| ITS Cluster 3:*Veillonella* | -0.382 (-2.606 to 1.843) | 1.114 | -0.343 | 66.695 | 0.733 |

**Table E15** Fixed effects estimates from a mixed-effects model of DLCO z-score, including ITS cluster, microbiota genera, and medication administration during hospitalisation. Interactions between independent variables are denoted by ':'.

| Independent Variable | Coefficient (95% CI) | Std. Error | Statistic | df | *P*-value |
|---|---|---|---|---|---|
| (Intercept) | -1.729 (-2.306 to -1.153) | 0.287 | -6.027 | 50.276 | 0.000 |
| polyn. shared *Veillonella* order:1 | -0.929 (-2.443 to 0.584) | 0.761 | -1.221 | 88.130 | 0.225 |
| polyn. shared *Veillonella* order:2 | 0.281 (-1.15 to 1.711) | 0.720 | 0.390 | 86.458 | 0.697 |
| Antibiotics (Yes) | 0.056 (-0.43 to 0.541) | 0.240 | 0.233 | 36.449 | 0.817 |
| Corticosteroids (Yes) | 0.315 (-0.103 to 0.732) | 0.207 | 1.522 | 39.849 | 0.136 |
| Inhaled Corticosteroids (Yes) | 0.553 (-0.479 to 1.586) | 0.512 | 1.080 | 43.754 | 0.286 |
| *Rothia*:ITS Cluster 1 | -1.539 (-3.574 to 0.496) | 1.024 | -1.503 | 89.038 | 0.136 |
| *Rothia*:ITS Cluster 2 | -1.835 (-3.739 to 0.069) | 0.959 | -1.914 | 90.634 | 0.059 |
| *Rothia*:ITS Cluster 3 | -7.123 (-14.154 to -0.091) | 3.533 | -2.016 | 79.107 | 0.047 |
| ITS Cluster 1:*Neisseria* | -1.267 (-3.239 to 0.705) | 0.992 | -1.276 | 89.634 | 0.205 |
| ITS Cluster 2:*Neisseria* | -0.274 (-2.896 to 2.349) | 1.320 | -0.207 | 89.859 | 0.836 |
| ITS Cluster 3:*Neisseria* | -1.245 (-3.227 to 0.736) | 0.996 | -1.250 | 83.322 | 0.215 |
| ITS Cluster 1:*Veillonella* | -0.927 (-2.807 to 0.953) | 0.947 | -0.979 | 91.985 | 0.330 |
| ITS Cluster 2:*Veillonella* | -3.672 (-5.955 to -1.388) | 1.149 | -3.195 | 90.123 | 0.002 |
| ITS Cluster 3:*Veillonella* | -0.76 (-3.127 to 1.607) | 1.191 | -0.638 | 86.077 | 0.525 |

**Table E16** Fixed effects estimates from a mixed-effects model of TLC z-score, including ITS cluster, microbiota genera, and medication administration during hospitalisation. Interactions between independent variables are denoted by ':'.

| **Independent Variable** | **Coefficient (95% CI)** | **Std. Error** | **Statistic** | **df** | ***P*-value** |
|---|---|---|---|---|---|
| (Intercept) | -1.726 (-2.47 to -0.982) | 0.367 | -4.699 | 37.340 | *0.000* |
| polyn. shared *Veillonella* order:1 | -1.687 (-3.181 to -0.193) | 0.748 | -2.256 | 63.273 | *0.028* |
| polyn. shared *Veillonella* order:2 | 0.459 (-0.861 to 1.778) | 0.660 | 0.695 | 60.451 | *0.490* |
| Antibiotics (Yes) | 0.044 (-0.717 to 0.804) | 0.375 | 0.116 | 35.633 | *0.908* |
| Corticosteroids (Yes) | -0.087 (-0.757 to 0.584) | 0.331 | -0.262 | 36.860 | *0.795* |
| Inhaled Corticosteroids (Yes) | 0.276 (-1.88 to 2.431) | 1.064 | 0.259 | 37.245 | *0.797* |
| *Neisseria*:ITS Cluster 1 | 0.768 (-1.496 to 3.031) | 1.133 | 0.678 | 63.736 | *0.500* |
| *Neisseria*:ITS Cluster 2 | -3.08 (-5.24 to -0.919) | 1.082 | -2.846 | 65.575 | *0.006* |
| *Neisseria*:ITS Cluster 3 | -0.409 (-2.169 to 1.352) | 0.883 | -0.463 | 68.901 | *0.645* |

**Table E17** Fixed effects estimates from a mixed-effects model of FEV1/FVC z-score, including ITS cluster, microbiota genera, and medication administration during hospitalisation. Interactions between independent variables are denoted by ':'.

| **Independent Variable** | **Coefficient (95% CI)** | **Std. Error** | **Statistic** | **df** | ***P*-value** |
|---|---|---|---|---|---|
| (Intercept) | 0.278 (-0.626 to 1.183) | 0.450 | 0.619 | 47.245 | 0.539 |
| polyn. shared *Veillonella* order:1 | 0.082 (-1.377 to 1.542) | 0.730 | 0.113 | 61.955 | 0.910 |
| polyn. shared *Veillonella* order:2 | -1.701 (-2.992 to -0.41) | 0.645 | -2.637 | 58.531 | 0.011 |
| Antibiotics (Yes) | 0.344 (-0.528 to 1.216) | 0.431 | 0.798 | 37.695 | 0.430 |
| Corticosteroids (Yes) | 0.003 (-0.744 to 0.75) | 0.369 | 0.009 | 39.411 | 0.993 |
| Inhaled Corticosteroids (Yes) | -1.11 (-3.605 to 1.386) | 1.234 | -0.899 | 39.180 | 0.374 |
| *Rothia*:ITS Cluster 1 | -0.61 (-2.923 to 1.703) | 1.161 | -0.525 | 75.535 | 0.601 |
| *Rothia*:ITS Cluster 2 | -1.268 (-3.435 to 0.9) | 1.088 | -1.165 | 75.990 | 0.248 |
| *Rothia*:ITS Cluster 3 | 6.662 (-0.076 to 13.401) | 3.372 | 1.976 | 62.806 | 0.053 |
| ITS Cluster 1:*Neisseria* | -0.329 (-2.376 to 1.718) | 1.025 | -0.321 | 65.107 | 0.749 |
| ITS Cluster 2:*Neisseria* | 1.761 (-0.546 to 4.067) | 1.156 | 1.523 | 67.051 | 0.132 |
| ITS Cluster 3:*Neisseria* | -0.731 (-2.657 to 1.194) | 0.963 | -0.760 | 60.548 | 0.450 |
| ITS Cluster 1:*Veillonella* | 1.151 (-0.842 to 3.145) | 0.999 | 1.153 | 66.804 | 0.253 |
| ITS Cluster 2:*Veillonella* | 0.92 (-1.376 to 3.217) | 1.151 | 0.800 | 68.930 | 0.427 |
| ITS Cluster 3:*Veillonella* | -1.277 (-3.688 to 1.134) | 1.208 | -1.058 | 65.735 | 0.294 |

**Table E18** Summary table for ITS cluster edge comparisons.

| Edges | Comparison | n | Statistic | *P*-value | *P*-adj |
|---|---|---|---|---|---|
| Inter-Kingdom Bact. and *Candida* | 1<->2 | 124 | 921.5 | 1.16e-06 | 1.62e-05 |
| Inter-Kingdom Bact. and *Candida* | 1<->3 | 167 | 3628.0 | 5.12e-01 | 1.00e+00 |
| Inter-Kingdom Bact. and *Candida* | 2<->3 | 141 | 3514.0 | 8.75e-09 | 1.31e-07 |
| Inter-Kingdom Bact. and *Cladosporium* | 1<->2 | 124 | 2043.0 | 2.91e-01 | 1.00e+00 |
| Inter-Kingdom Bact. and *Cladosporium* | 1<->3 | 167 | 1263.5 | 1.68e-12 | 2.86e-11 |
| Inter-Kingdom Bact. and *Cladosporium* | 2<->3 | 141 | 795.5 | 2.29e-10 | 3.66e-09 |
| Inter-Kingdom | 1<->2 | 124 | 2070.0 | 2.35e-01 | 1.00e+00 |
| Inter-Kingdom | 1<->3 | 167 | 2829.5 | 4.60e-02 | 3.68e-01 |
| Inter-Kingdom | 2<->3 | 141 | 1641.0 | 8.00e-03 | 7.20e-02 |
| Inter-Kingdom Bact. and *Saccharomyces* | 1<->2 | 124 | 3258.5 | 3.20e-13 | 5.76e-12 |
| Inter-Kingdom Bact. and *Saccharomyces* | 1<->3 | 167 | 4873.0 | 4.32e-06 | 5.62e-05 |
| Inter-Kingdom Bact. and *Saccharomyces* | 2<->3 | 141 | 1294.5 | 3.05e-05 | 3.35e-04 |
| Bacterial Intra-Kingdom | 1<->2 | 124 | 2028.0 | 3.31e-01 | 1.00e+00 |
| Bacterial Intra-Kingdom | 1<->3 | 167 | 3267.5 | 5.58e-01 | 1.00e+00 |
| Bacterial Intra-Kingdom | 2<->3 | 141 | 1875.0 | 1.01e-01 | 7.07e-01 |
| Fungal Intra-Kingdom | 1<->2 | 124 | 2695.5 | 1.17e-05 | 1.40e-04 |
| Fungal Intra-Kingdom | 1<->3 | 167 | 3656.0 | 5.08e-01 | 1.00e+00 |
| Fungal Intra-Kingdom | 2<->3 | 141 | 1347.0 | 8.67e-05 | 8.67e-04 |

**Table E19** Summary table for mean and standard deviation (sd) of ITS cluster edge statistics.

| **Edges** | **ITS Cluster** | **mean** | **sd** |
|---|---|---|---|
| Inter-Kingdom Bact. and *Candida* | 1 | 2.41 | 3.21 |
| Inter-Kingdom Bact. and *Candida* | 2 | 5.47 | 2.94 |
| Inter-Kingdom Bact. and *Candida* | 3 | 2.07 | 3.15 |
| Inter-Kingdom Bact. and *Cladosporium* | 1 | 4.48 | 2.93 |
| Inter-Kingdom Bact. and *Cladosporium* | 2 | 3.98 | 3.36 |
| Inter-Kingdom Bact. and *Cladosporium* | 3 | 8.36 | 3.08 |
| Inter-Kingdom | 1 | 23.08 | 7.23 |
| Inter-Kingdom | 2 | 21.29 | 9.10 |
| Inter-Kingdom | 3 | 25.52 | 8.33 |
| Inter-Kingdom Bact. and *Saccharomyces* | 1 | 9.59 | 2.50 |
| Inter-Kingdom Bact. and *Saccharomyces* | 2 | 4.27 | 3.35 |
| Inter-Kingdom Bact. and *Saccharomyces* | 3 | 7.10 | 3.72 |
| Bacterial Intra-Kingdom | 1 | 776.91 | 229.31 |
| Bacterial Intra-Kingdom | 2 | 697.12 | 317.22 |
| Bacterial Intra-Kingdom | 3 | 797.23 | 276.33 |
| Fungal Intra-Kingdom | 1 | 417.29 | 82.54 |
| Fungal Intra-Kingdom | 2 | 324.65 | 114.95 |
| Fungal Intra-Kingdom | 3 | 405.55 | 96.91 |

#### Supplementary Figure legends

**Supplementary Figure E1** Overview of sample availability. Dumbbell plot representing sample availability (oropharyngeal and/or rectal swabs) per patient, over the study period. Each line represents one patient. Yellow = available oropharyngeal swab, green = both oropharyngeal and rectal swabs available. T1 = 3 -months, T2 = 6 -months, T3 = 12 -months post-hospitalisation.

**Supplementary Figure E2** **(A)** URT 16S load over the study period. T1 = 3 -months, T2 = 6 -months, T3 = 12 -months post-hospitalization. Comparisons were made using Kruskal-Wallis with Dunn's post-hoc test and Bonferroni adjustment. **(B)** Gut 16S load over the study period. T1 = 3 -months, T2 = 6 -months, T3 = 12 -months post-hospitalization. Comparisons were made using Kruskal-Wallis with Dunn's post-hoc test and Bonferroni adjustment.

**Supplementary Figure E3** Dynamics in lung function metrics (TLC, FVC, FEV1 and DLCO z-scores) per COVID-19 -related AHFR-BLI status, over the study period. Each dot represents one sample. T1 = 3 -months, T2 = 6 -months, T3 = 12 -months post-hospitalization. Comparisons were made using Wilcoxon-test.

**Supplementary Figure E4** **(A)** 16S load over the study period per COVID-19 -related AHFR-BLI status. T1 = 3 -months, T2 = 6 -months, T3 = 12 -months post-hospitalization. URT (left) and gut (right). Comparisons were made using Wilcoxon-test. **(B)** Alpha diversity dynamics (Chao1 and Shannon) over the study period relative to antibiotic administration during hospitalization. Each dot represents one sample. T1 = 3 -months, T2 = 6 -months, T3 = 12 -months post-hospitalization. URT (left) and gut (right). Comparisons were made using Wilcoxon-test. *P-value significance: *. < 0.05, **. ≤ 0.01, ***. ≤ 0.001.* **(C)** URT alpha diversity dynamics (Chao1 and Shannon) over the study period relative to intubation status during hospitalization. Each dot represents one sample. T1 = 3 -months, T2 = 6 -months, T3 = 12 -months post-hospitalization. Comparisons were made using Wilcoxon-test. **(D)** Alpha diversity dynamics (Chao1 and Shannon) over the study period among post- AHFR-BLI subjects, relative to intubation status during hospitalization. Each dot represents one sample. T1 = 3 -months, T2 = 6 -months, T3 = 12 -months post-hospitalization. Comparisons were made using Wilcoxon-test. **(E)** Gut alpha diversity dynamics (Chao1 and Shannon) over the study period relative to intubation status during hospitalization. Each dot represents one sample. T1 = 3 -months, T2 = 6 -months, T3 = 12 -months post-hospitalization. Comparisons were made using Wilcoxon-test.

**Supplementary Figure E5** Dumbbell plot representing lung function recovery trajectories over the study period, per pulmonary function metric (TLC, FVC, DLCO and FEV1). Each line represents one patient. T1 = 3 -months, T2 = 6 -months, T3 = 12 -months post-hospitalization.

**Supplementary Figure E6** (**A**) Stacked bar plots representing the relative abundance of the top 20 genera in the URT per lung function recovery trajectory (TLC, FCV, DLCO and FEV1). The "other" category summarizes all remaining genera. N = number of samples per group. (**B**) Stacked bar plots representing the relative abundance of the top 20 genera in the gut per lung function recovery trajectory (TLC, FCV, DLCO and FEV1). The "other" category summarizes all remaining genera. N = number of samples per group.

**Supplementary Figure E7** Non-linear relationship between gut-lung shared *Veillonella* and lung function. Scatter plots representing the relationship between gut-lung shared *Veillonella* abundance and **(A)** FVC or **(B)** FEV1. The blue line represents a second-order orthogonal polynomial, and in grey the 95% confidence interval.

**Supplementary Figure E8** **(A)** ITS alpha diversity dynamics (Chao1 and Shannon) over the study period. Each dot represents one sample. T1 = 3 -months, T2 = 6 -months, T3 = 12 -months post-hospitalization. Comparisons were made using Kruskal-Wallis with Dunn's post-hoc test and Bonferroni adjustment. **(B)** URT ITS load over the study period. T1 = 3 -months, T2 = 6 -months, T3 = 12 -months post-hospitalization. Each dot represents one sample. Comparisons were made using Kruskal-Wallis with Dunn's post-hoc test and Bonferroni adjustment. **(C)** URT ITS load per ITS cluster. Each dot represents one sample. Comparisons were made using Kruskal-Wallis with Dunn's post-hoc test and Bonferroni adjustment. *P-value significance: *. < 0.05, **. ≤ 0.01, ***. ≤ 0.001.*

### Figure E1

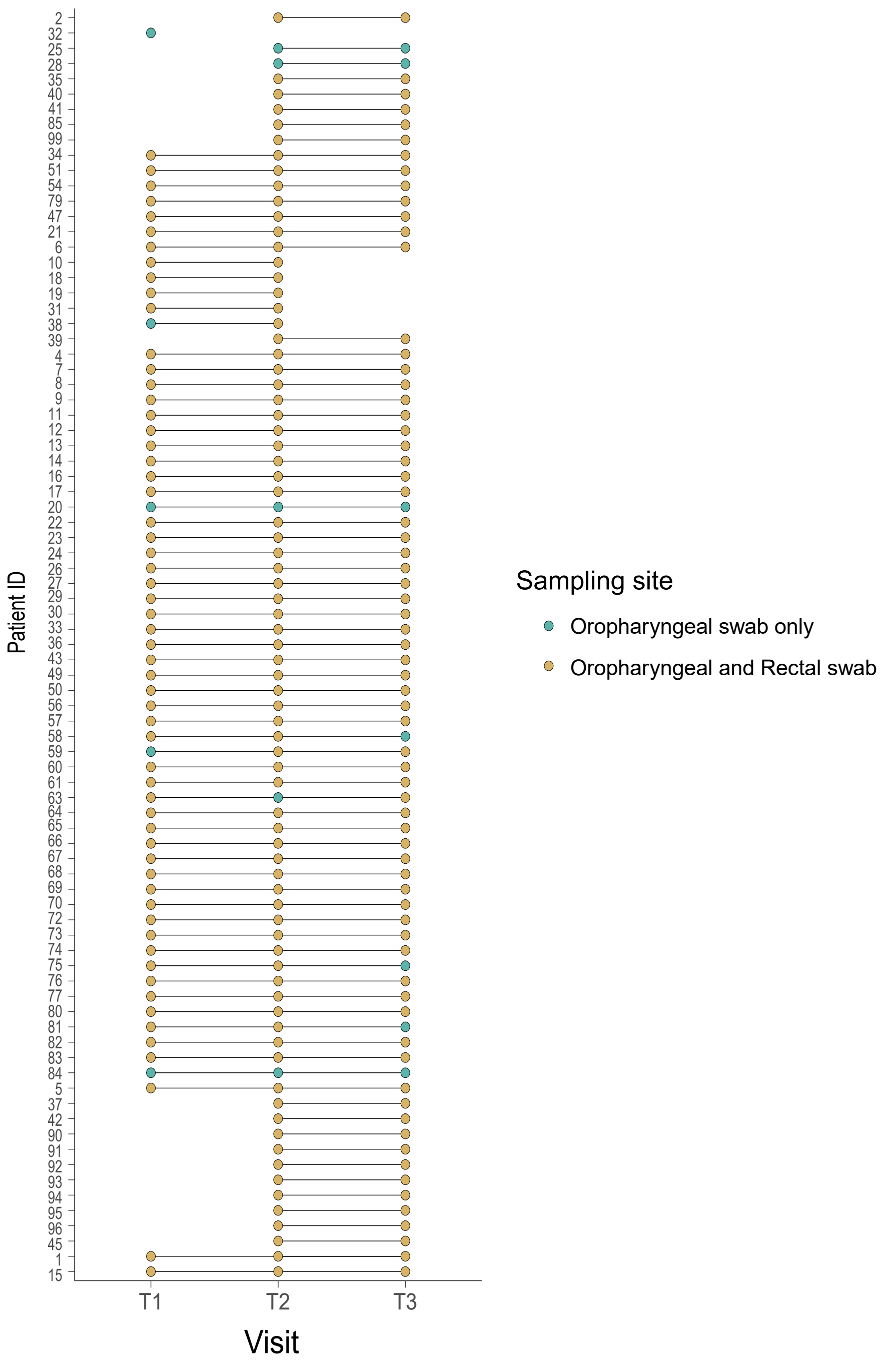

Patient ID
2
32
25
28
35
40
41
85
99
34
51
54
79
47
21
6
10
18
19
31
38
39
4
7
8
9
11
12
13
14
16
17
20
22
23
24
26
27
29
30
33
36
43
49
50
56
57
58
59
60
61
63
64
65
66
67
68
69
70
72
73
74
75
76
77
80
81
82
83
84
5
37
42
90
91
92
93
94
95
96
45
1
15
T1
T2
T3
Visit
Sampling site
Oropharyngeal swab only
Oropharyngeal and Rectal swab

Figure E2

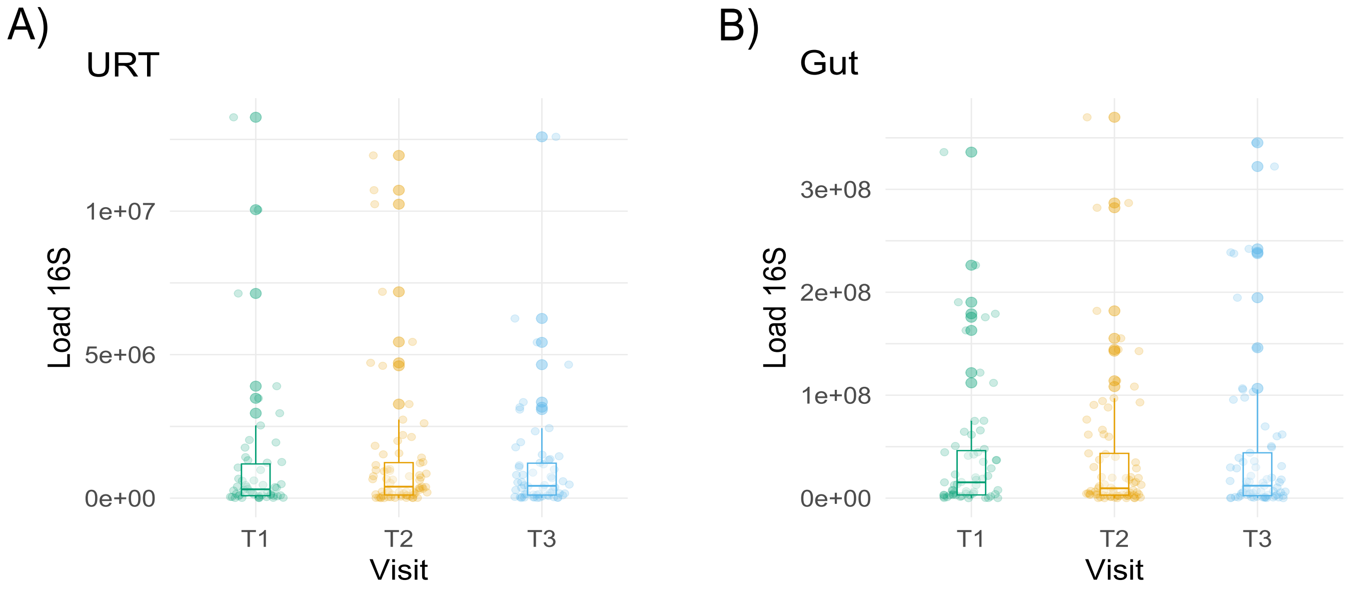

A)
URT
Load 16S
1e+07
5e+06
0e+00
T1
T2
T3
Visit
B)
Gut
Load 16S
3e+08
2e+08
1e+08
0e+00
T1
T2
T3
Visit

### Figure E 3

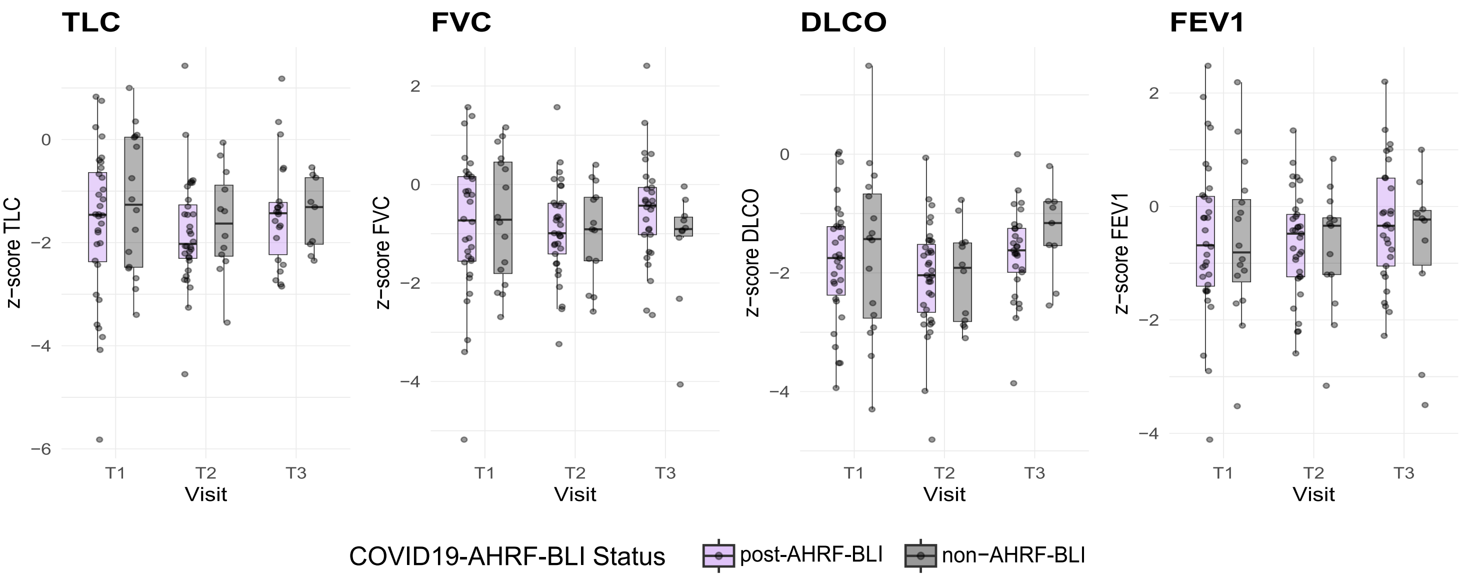
TLC
FVC
DLCO
FEV1
z−score TLC
z−score FVC
z−score DLCO
z−score FEV1
Visit
T1
T2
T3
COVID19-AHRF-BLI Status
post-AHRF-BLI
non−AHRF-BLI

### Figure E4

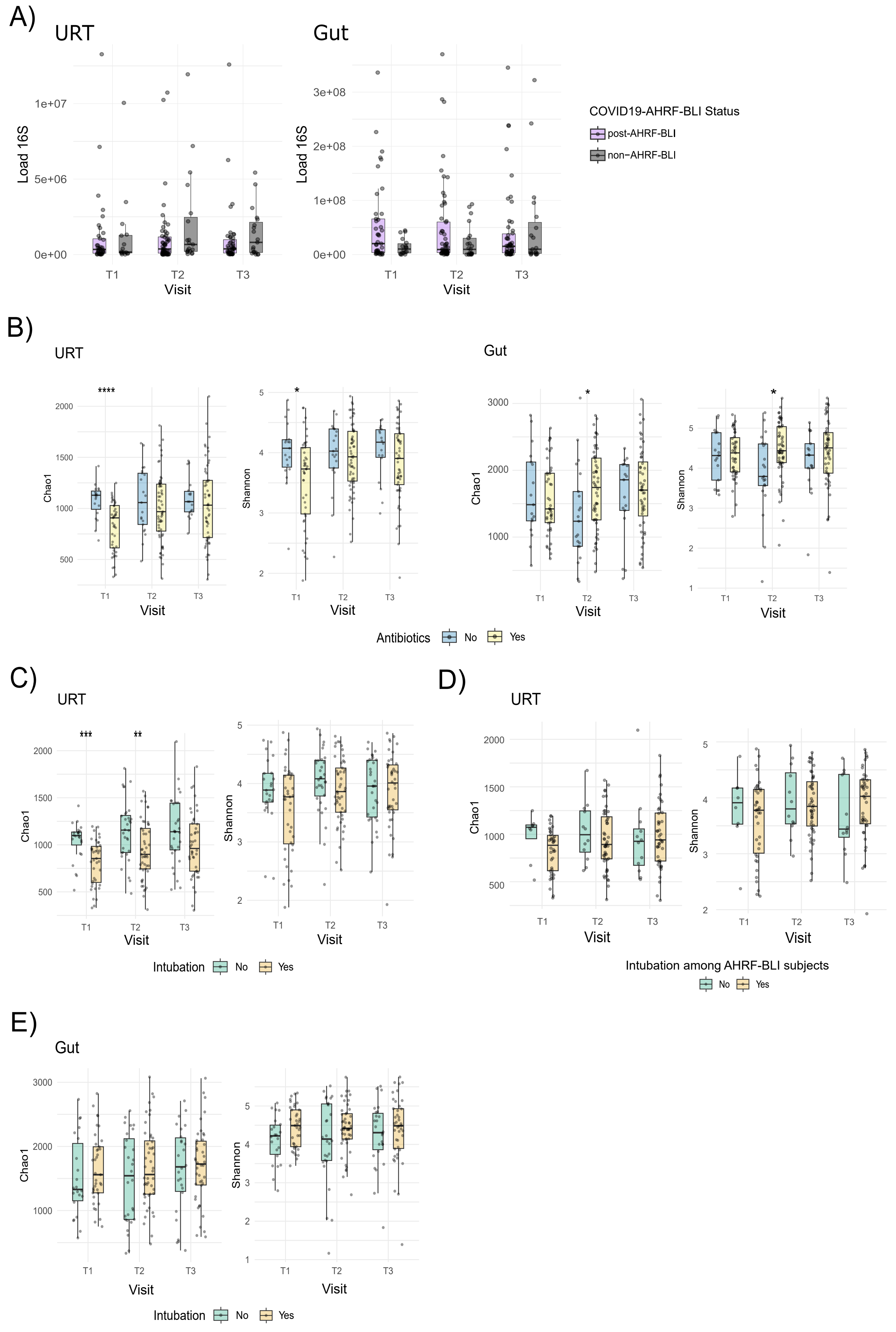
A)
URT
Gut
Load 16S
Visit
T1
T2
T3
COVID19-AHRF-BLI Status
post-AHRF-BLI
non−AHRF-BLI
B)
URT
Gut
Chao1
Shannon
****
*
Antibiotics
No
Yes
C)
URT
***
**
Intubation
No
Yes
D)
URT
Intubation among AHRF-BLI subjects
No
Yes
E)
Gut
Intubation
No
Yes

### Figure E5

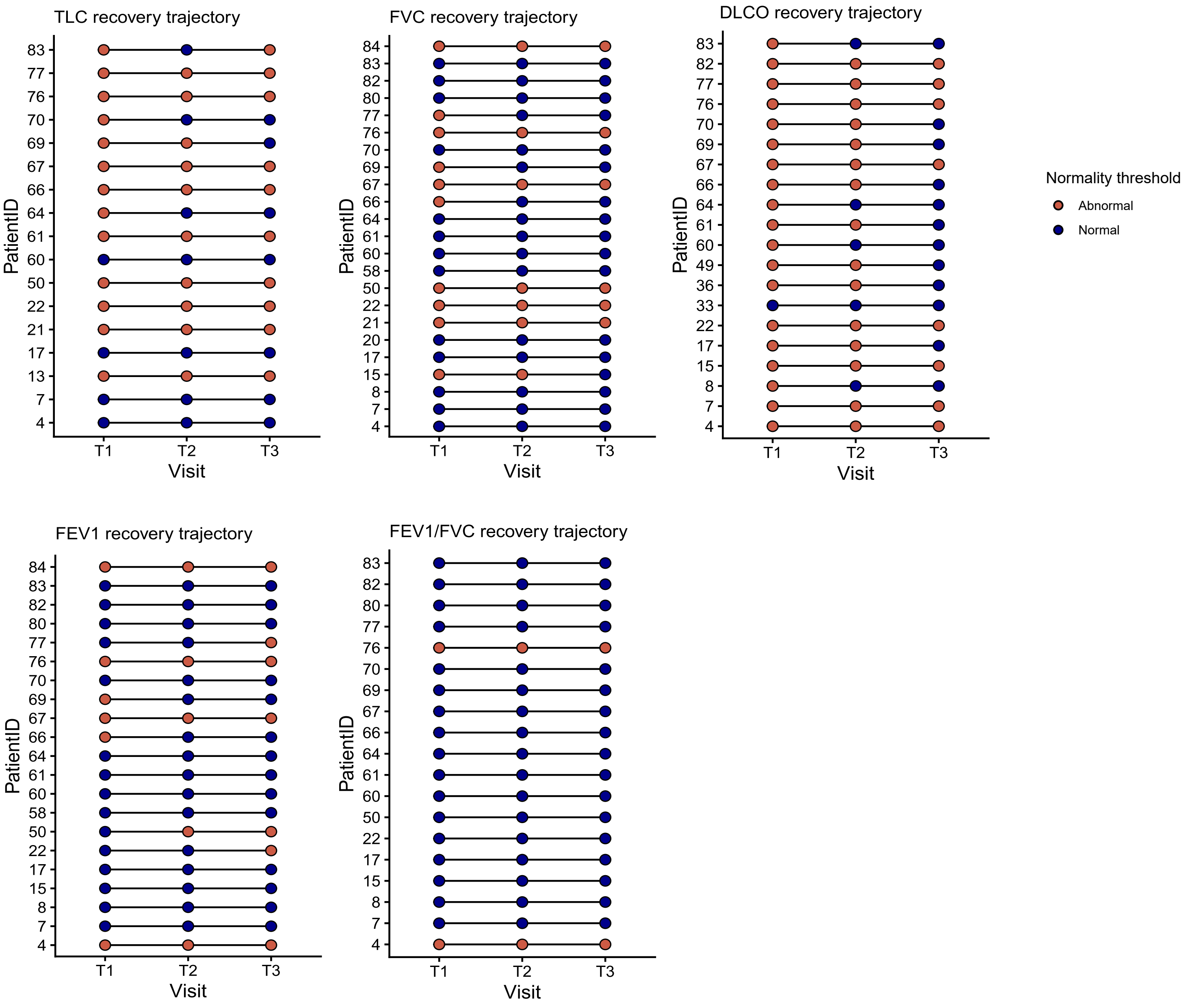

TLC recovery trajectory
FVC recovery trajectory
DLCO recovery trajectory
FEV1 recovery trajectory
FEV1/FVC recovery trajectory
PatientID
Visit
T1
T2
T3
Normality threshold
Abnormal
Normal

### Figure E6

#### A) URT

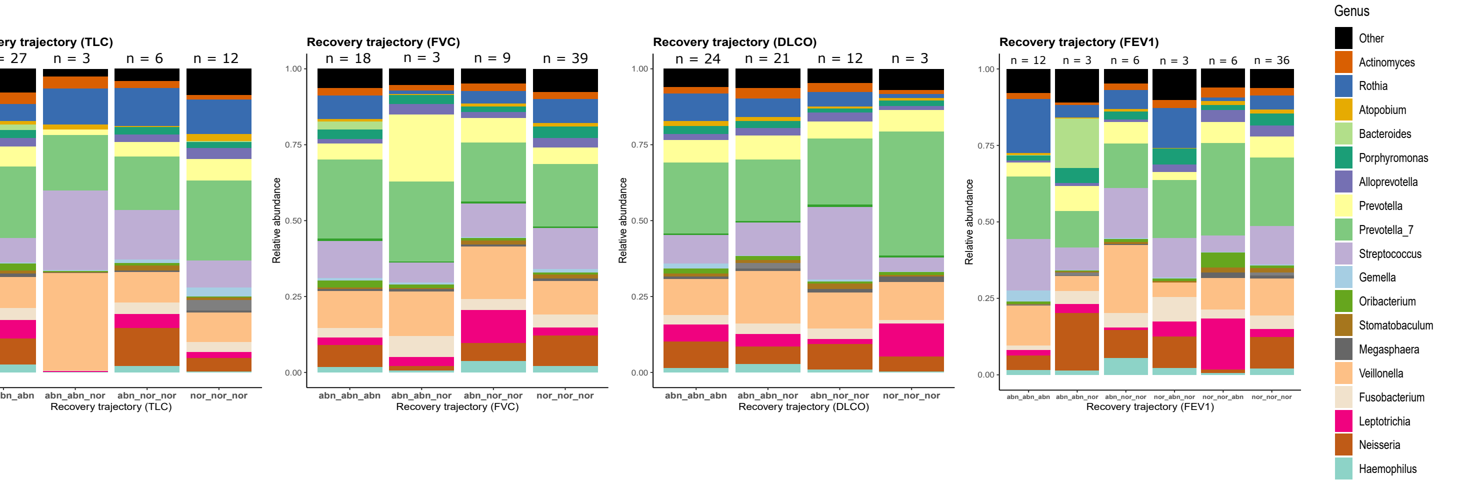


#### B) Gut

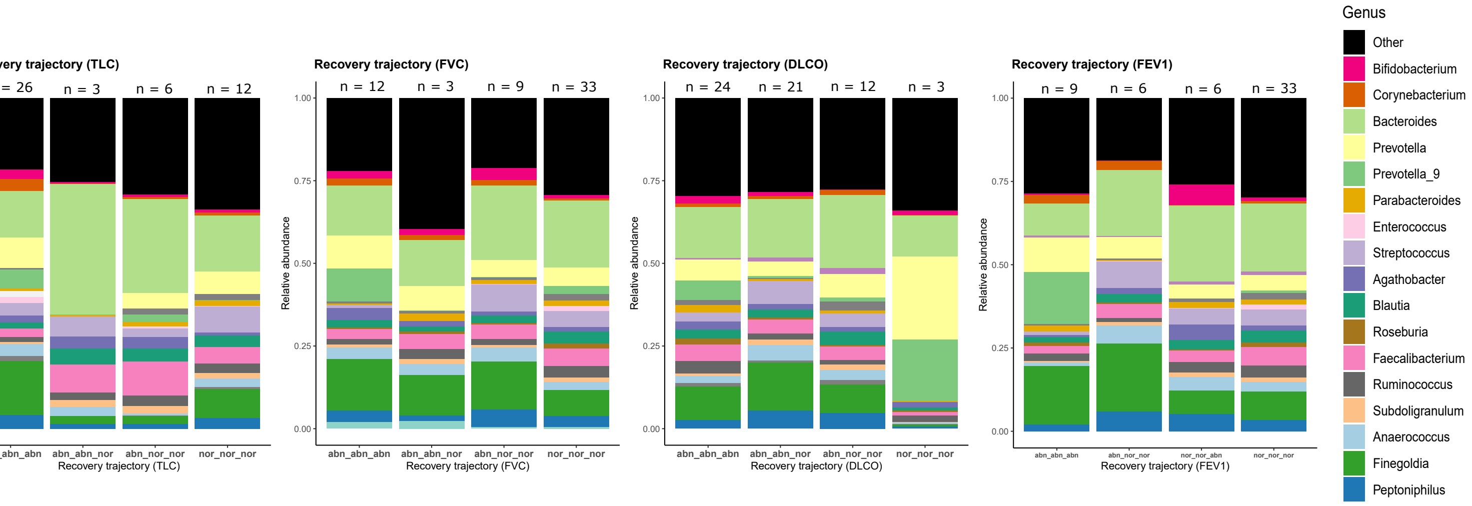

Figure E7

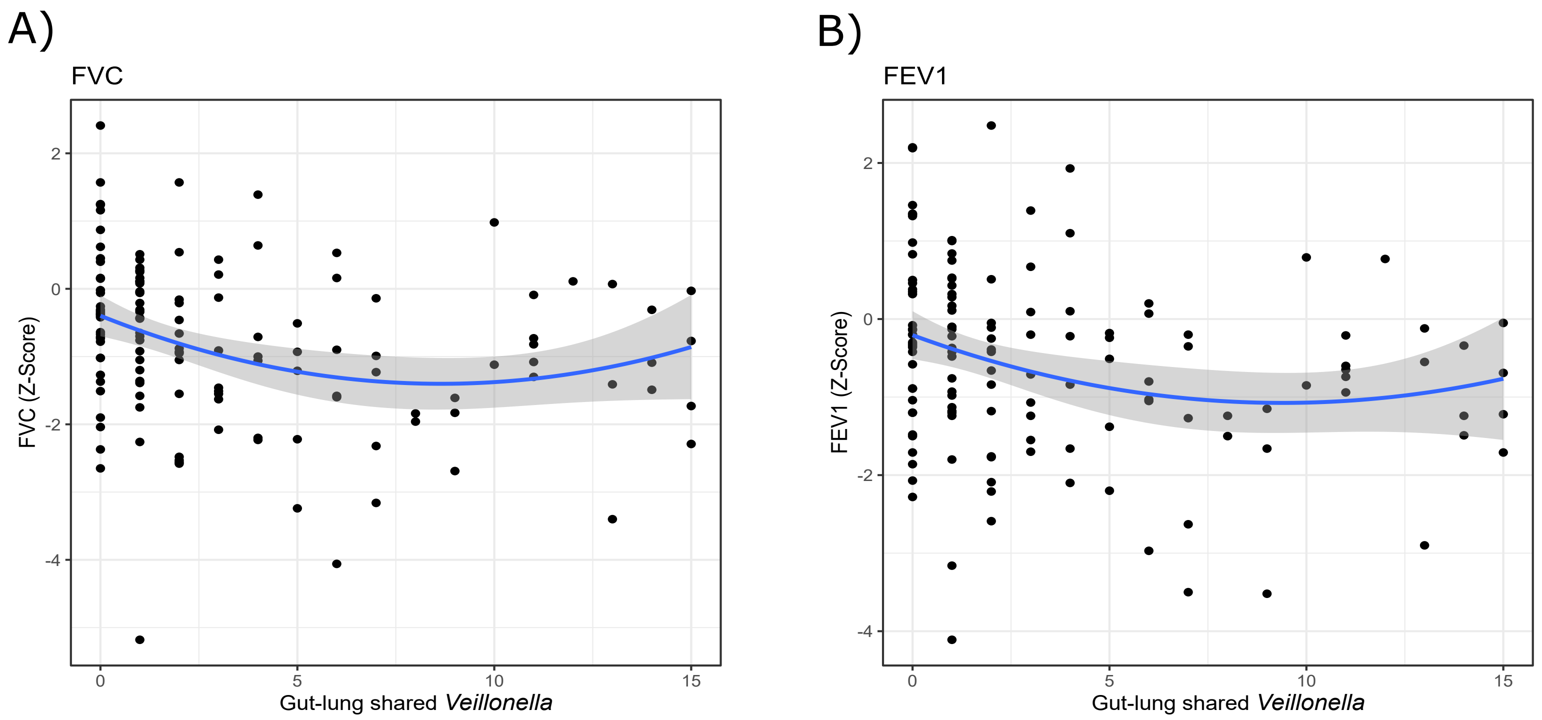

A)
FVC
FVC (Z-Score)
2
0
-2
-4
0
5
10
15
Gut-lung shared *Veillonella*
B)
FEV1
FEV1 (Z-Score)
2
0
-2
-4
0
5
10
15
Gut-lung shared *Veillonella*

### Figure E8

A)

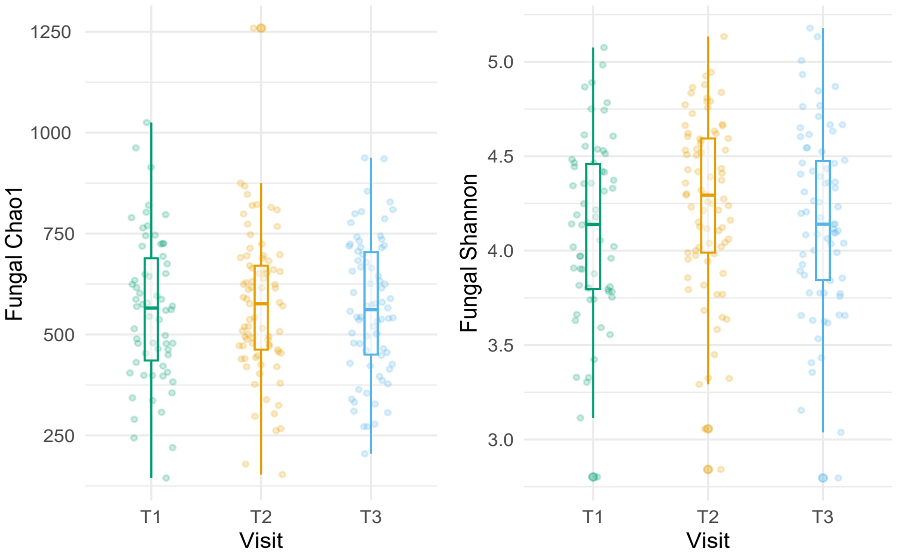


B)

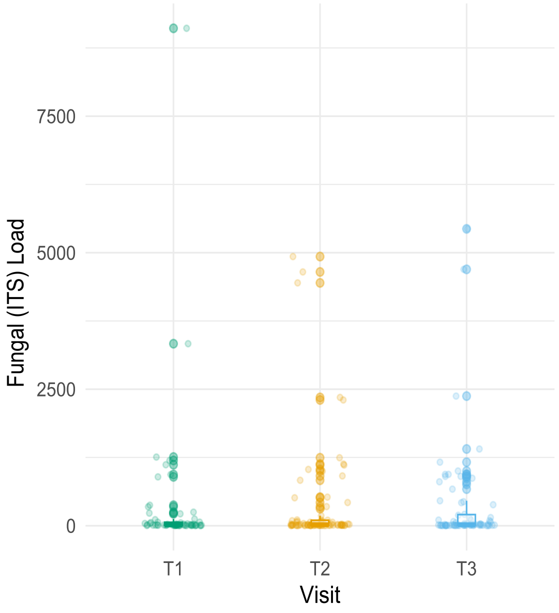


C)

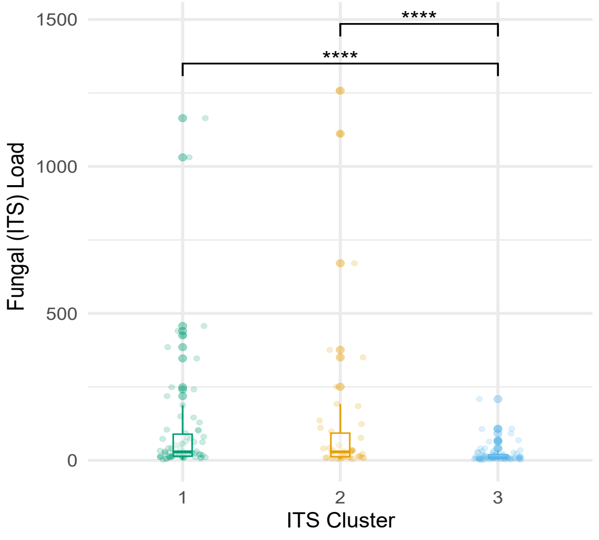
